# Influenza virus shedding and symptoms: Dynamics and implications from a multi-season household transmission study

**DOI:** 10.1101/2024.03.04.24303692

**Authors:** Sinead E. Morris, Huong Q. Nguyen, Carlos G. Grijalva, Kayla E. Hanson, Yuwei Zhu, Jessica E. Biddle, Jennifer K. Meece, Natasha B. Halasa, James D. Chappell, Alexandra M. Mellis, Carrie Reed, Matthew Biggerstaff, Edward A. Belongia, H. Keipp Talbot, Melissa A. Rolfes

**Author notes:** **Corresponding author**: Sinead Morris, Centers for Disease Control and Prevention, 1600 Clifton Rd NE, Atlanta, GA, 30033. **Disclaimer:** The findings and conclusions in this report are those of the authors and do not necessarily represent the views of the Centers for Disease Control and Prevention. **Competing Interest Statement:** H.Q.N receives research support unrelated to this work from CSL Seqirus and GSK, and honorarium for participating in a consultancy group for Moderna outside the submitted work. C.G.G received research support unrelated to this work from Merck. K.E.H, J.K.M, and E.A.B receive research support unrelated to this work from CSL Seqirus. The authors have no competing interests to declare.

## Abstract

Isolation of symptomatic infectious persons can reduce influenza transmission. However, virus shedding that occurs without symptoms will be unaffected by such measures. Identifying effective isolation strategies for influenza requires understanding the interplay between individual virus shedding and symptom presentation. From 2017–2020, we conducted a case-ascertained household transmission study using influenza real-time reverse transcription quantitative PCR (RT-qPCR) testing of nasal swabs and daily symptom diary reporting for up to 7 days after enrollment (≤14 days after index onset). We assumed real-time RT-qPCR cycle threshold (Ct) values were indicators of quantitative virus shedding and used symptom diaries to create a score that tracked influenza-like-illness (ILI) symptoms (fever, cough, or sore throat). We fit phenomenological nonlinear mixed-effects models stratified by age and vaccination status and estimated two quantities influencing isolation effectiveness: shedding before symptom onset and shedding that might occur once isolation ends. We considered different isolation end points (including 24 hours after fever resolution or 4 days after symptom onset) and assumptions about the infectiousness of Ct shedding trajectories. Of the 116 household contacts with ≥2 positive tests for longitudinal analyses, 105 (91%) experienced ≥1 ILI symptom. On average, children <5 years experienced greater peak shedding, longer durations of shedding, and elevated ILI symptom scores compared with other age groups. Most individuals (63/105) shed <10% of their total shed virus before symptom onset, and shedding after isolation varied substantially across individuals, isolation end points, and infectiousness assumptions. Our results can inform strategies to reduce transmission from symptomatic individuals infected with influenza.

**Significance Statement:** Individuals infected with influenza are encouraged to avoid contact with others for a period following symptom onset. This action should reduce the likelihood of onward transmission if infectious virus shedding is associated with symptom presentation. We modeled influenza virus shedding and symptom dynamics in participants of a multi-season household transmission study. On average, children <5 years shed more virus for longer durations and experienced elevated influenza-like-illness symptoms compared with older age groups. Most shedding took place after symptom onset, and estimated shedding that might remain after a period of avoiding contact with others depended on how the end of this period was defined. Our results can help inform strategies to reduce transmission from symptomatic individuals infected with influenza.

## Introduction

Seasonal influenza causes substantial morbidity and mortality in the United States each year [1]. Measures to mitigate this burden include pharmaceutical interventions, such as vaccination, and nonpharmaceutical interventions, such as case isolation. The latter is designed to reduce exposure to virus shedding from symptomatic individuals, thereby reducing the potential for onward transmission. The Centers for Disease Control and Prevention (CDC) recommends symptomatic individuals stay at home and avoid contact with others until 24 hours after fever resolution, or for 4-5 days following symptom onset if they do not experience fever [2, 3]. However, transmission will be unaffected by such mitigation measures if it occurs in the absence of symptoms (for example, prior to symptom onset or from asymptomatic individuals) or after the recommended duration has passed. Understanding how the timing and progression of influenza symptoms relate to influenza virus shedding and transmission is crucial to determining the potential impact of different strategies for reducing onward transmission from symptomatic individuals [4, 5].

Studies of influenza transmission within households provide valuable information on the natural history of infection [6]. Individual shedding and infection progression can be followed through routine PCR testing of household members and recording of symptoms that arise [7, 8]. Such information can be used to characterize patterns of virus shedding from asymptomatic and symptomatic individuals, and to estimate household secondary attack rates and influenza vaccine effectiveness [9–13]. Individual-level data can also be modeled to explore potential differences by age or vaccination status that may impact the effectiveness of isolation [14–18]. Such insights have improved our understanding of effective isolation measures for SARS-CoV-2 but are limited for influenza [19–21].

Here we modeled individual-level data from a household transmission study conducted across three influenza seasons in the United States. We used cycle threshold (Ct) values obtained from real-time reverse transcription quantitative PCR (RT-qPCR) testing of individual nasal swab specimens to infer longitudinal virus shedding dynamics, and used symptom diaries to assess the relative timing and progression of influenza-associated symptoms. We then identified differences in shedding and symptom presentation by age and vaccination status, and estimated levels of pre-symptomatic shedding and shedding that may remain once a mitigation measure, such as isolation, has ended. Our findings can help inform effective strategies to reduce transmission from symptomatic individuals infected with influenza.

## Materials and Methods

### Study

We conducted a case-ascertained household transmission study during the 2017-2018, 2018-2019 and 2019-2020 influenza seasons. Households were enrolled following the recruitment of index cases with laboratory-confirmed influenza virus infection from clinics or testing sites in Wisconsin and Tennessee. Households enrolled within seven days of the index illness onset date were eligible to participate and followed for up to seven days after enrollment. Participants provided demographic information including age and vaccination status at enrollment. Written informed consent was obtained for all participants. Study protocols and procedures were reviewed by the Institutional Review Boards at the Marshfield Clinic Research Institute and Vanderbilt University. CDC determined these activities were conducted consistent with applicable federal law and CDC policy (see 45 C.F.R. part 46; 21 C.F.R. part 56). The study has been described in detail elsewhere [9, 22]; information relevant for the current analysis is summarized below.

### Virus shedding

Nasal swabs were collected daily from participants (either by study staff or self-/adult-collected) and tested for influenza by real-time RT-qPCR. Testing was performed using the CDC Human Influenza Virus Real-Time RT-PCR Diagnostic Panel and Influenza A/B Typing Kit at Marshfield Clinic Research Institute or Vanderbilt University Medical Center. We inferred that real-time RT-qPCR Ct values were an indicator of virus shedding and hereafter refer to samples with PCR positive Ct values as reflecting shed virus [23]. Since a positive Ct value does not necessarily reflect infectious virus, our Ct analyses relate to the total viral genomic material an individual may shed, not solely that which is infectious. We explore different assumptions about the relationship between Ct measurements and infectious virus in our analyses below.

Ct values are inversely proportional to viral load such that higher Ct values reflect lower amounts of viral material in a sample. Forty-five cycles were performed for each reaction and samples with Ct values <40 were deemed positive [24, 25]. Positive samples with values approaching 40 (e.g., values >36) were repeated to ensure results were reproducible and all negative tests were assigned a value of 40. When individuals had a self- and staff-collected test result on the same day, the Ct values were significantly correlated (correlation = 0.6, p<0.0001) and so we used the result from the self-collected swab for consistency. Although Ct values can vary between laboratories, we found no evidence of systematic differences in Ct dynamics by testing site (Figure S1) and thus combined data from both sites in all analyses.

### Symptom scores

Symptom diaries identifying the presence of eight possible signs and symptoms associated with influenza infection (fever, cough, sore throat, runny nose, nasal congestion, fatigue, shortness of breath, and wheezing) were self-/adult-administered daily (Figure 1A). We translated these diaries into a quantitative score, S_ILI_, that measured the sum of influenza-like-illness (ILI) signs or symptoms reported on each day as

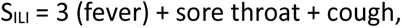

**Figure 1.**
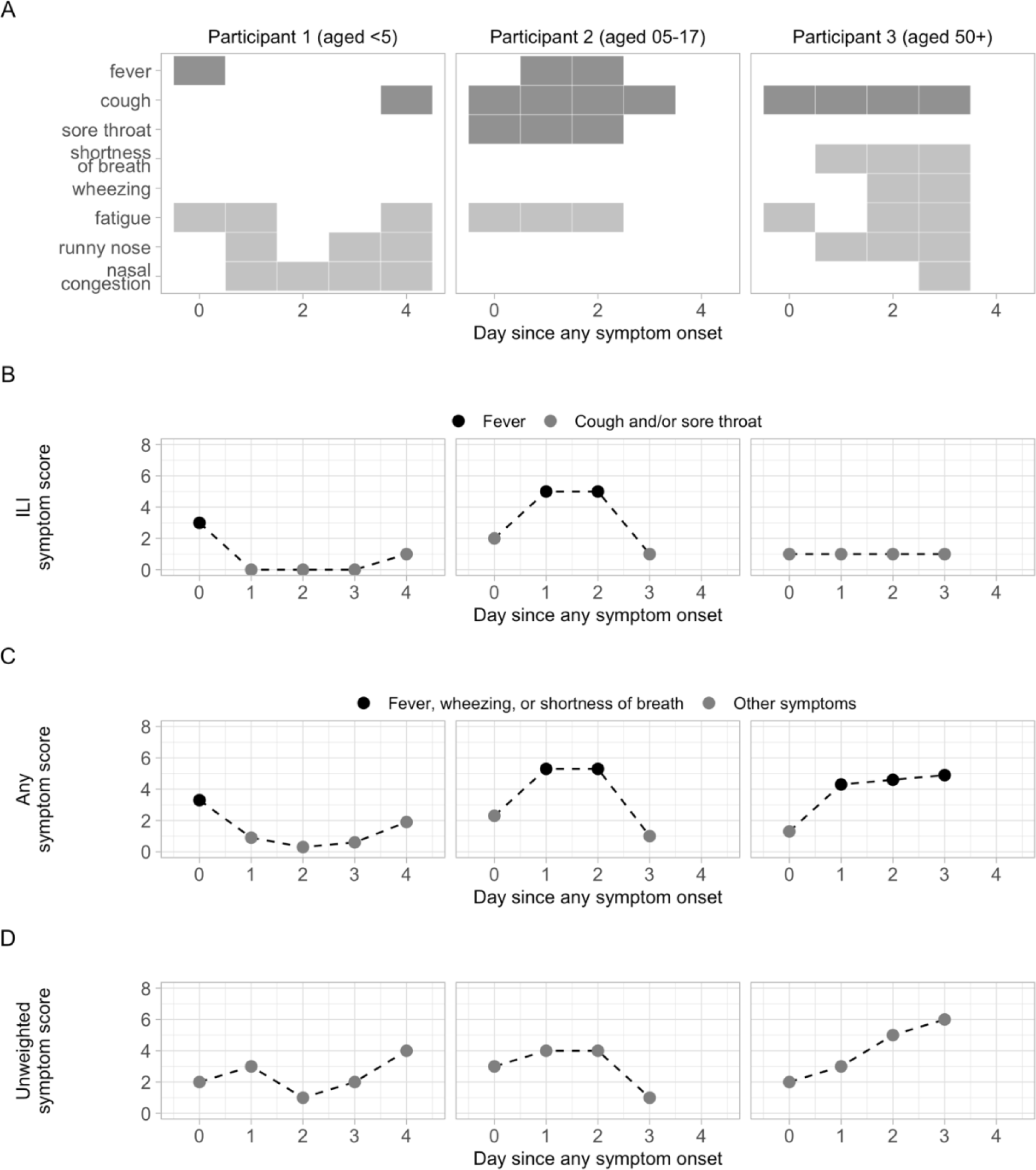
Translation of symptom diaries to symptom scores. (A) Symptom diary reports from a child aged <5 years (left panel), a child aged 5-17 (middle) and an adult aged 50+ (right). ILI signs or symptoms (fever, cough, sore throat) reported on a given day are highlighted in dark grey; all other reported symptoms are shown in light grey. Diary reports from (A) are translated to ILI symptom scores, S_ILI_ (B), any symptom scores, S_ANY_ (C), and unweighted symptom scores, S_UNW_ (D). In (B) and (C), colors indicate the presence of fever (B), or fever, wheezing, or shortness of breath (C). Note these colors have different interpretations to those in (A). ILI stands for influenza-like-illness.

where fever, sore throat, and cough equal 1 on days they were reported and 0 on days they were not (Figure 1B). Fever was weighted more heavily as a proxy for increased clinical severity [26–28]. We also defined an alternative score, S_ANY_, that accounted for any sign or symptom assessed in the diaries. In this score, shortness of breath and wheezing were more heavily weighted, in addition to fever, to include greater lower respiratory tract involvement as a proxy for severity (Figure 1C) [26, 27, 29]. A value of 3 was assigned if at least one of fever, shortness of breath, or wheezing were reported; this value did not increase if more than one was reported. Additional symptoms that were not included in the ILI definition (nasal congestion, runny nose, and fatigue) were each weighted by 1/3, so that

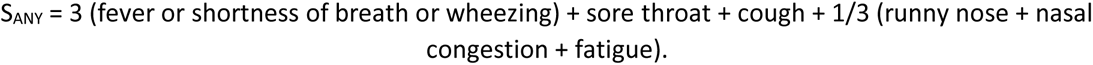

The weights for S_ILI_ and S_ANY_ were chosen to allow for back-translation of scores to the presence or absence of fever, shortness of breath, or wheezing. For example, S_ILI_ > 2 can only occur if fever is reported and 2 ≥ S_ILI_ > 1 can only occur if cough and sore throat are reported without fever. Similarly, S_ANY_ > 3 can only occur if fever, shortness of breath, or wheezing are among the reported symptoms; this would not be the case if runny nose, nasal congestion, or fatigue were weighted by 1 instead of 1/3. Finally, we defined an unweighted score, S_UNW_, that was simply the number of signs or symptoms reported on any given day [10]. We compared this with S_ILI_ and S_ANY_ to assess the sensitivity of our results to the choice of scoring weights.

### Model

We assume viral shedding over time within an infected individual is proportional to a continuous probability distribution, f_v_(t, a_v_, b_v_). Here, a_v_ and b_v_ are the distribution parameters, and t represents days since ILI symptom onset (for analysis of individuals reporting ILI symptoms); days since any symptom onset (for analysis of individuals reporting any symptoms); or days since first positive test (for analysis of asymptomatic individuals). To allow variation in the magnitude and timing of shedding, we model the amount of virus shed at time t, V(t), by

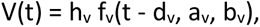

where h_v_ > 0 is a magnitude scaling factor and d_v_ a shift in time. Similarly, we model the symptom score (S_ILI_, S_ANY_, or S_UNW_) of a symptomatic infected individual as

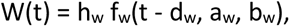

where t represents days since ILI symptom onset (for S_ILI_) or days since any symptom onset (for S_ANY_ and S_UNW_). We investigate three plausible distributions for f_v_ and f_w_: lognormal, gamma, and Weibull distributions [4, 11, 19, 30]. Since Ct values are bounded by 40 with higher values reflecting lower levels of detected virus, we fit the Ct data with a transformed function, ^*V̂*^(t) = 40 – V(t).

### Model fitting and covariate analysis

Household contacts with at least two Ct values <40 were included in the modeling analysis. To mitigate the impact of symptoms that were consistently reported due to causes other than influenza, we discarded all data from individuals who reported symptoms on three or more days before testing positive, when those days also coincided with a negative test. For analysis of symptomatic infection, we included individuals whose first swab (positive or negative) was on or before symptom onset, and t = 0 anchored all trajectories at symptom onset. For analysis of asymptomatic infection, we could not use time since symptom onset as a common anchor point. We therefore only included individuals with ‘incident’ infection (i.e., those with a recorded negative test before their first positive test). This allowed us to use time since first positive test as a common anchor point that was at, or near, the onset of virus shedding (Figure S2).

We fit ^*V̂*^(t) and W(t) to the Ct and symptom score data, respectively, using a nonlinear mixed effects framework [31, 32]. We assumed both variables were normally-distributed with constant error terms, and ensured model residuals were normally-distributed using the Shapiro-Wilk test [33]. Each parameter was assigned a fixed effect and an individual-level random effect; the latter captured variation between participants. We assumed a, b and h were lognormally distributed to ensure positivity, and d was normally distributed to allow positive or negative shifts in time. All parameters were assumed to be independent, and we explored models that stratified one or more parameters by candidate covariates for age group, vaccination status, virus type and season. The importance of each covariate-parameter relationship was evaluated using ANOVA [31, 32].

The fit of each candidate model (lognormal, gamma, or Weibull) was compared using Akaike Information Criterion (AIC), where lower AIC values reflect greater statistical support [34]. For the model with lowest AIC, we simulated Ct and symptom trajectories for each individual using the fitted parameter estimates. We then calculated key summary metrics, including the duration of shedding (or duration of symptoms), the timing of peak shedding (or timing of peak symptom scores), and the area under each curve. We tested for associations between these summary metrics and each candidate covariate (listed above) using the non-parametric Kruskal-Wallis test and controlled the false-discovery rate using the Benjamini-Hochberg correction. Further details on summary metric calculations are provided in the Supplementary Information.

Due to sample size, we did not perform additional stratifications by multiple covariates when testing for associations with the summary metrics described above. Instead, to distinguish covariates with the greatest independent influence on trajectory dynamics from those acting primarily through collinearity with other covariates, we performed hierarchical partitioning [35, 36]. Hierarchical partitioning provides a systematic way to assess the independent influence of a particular covariate on a dependent variable, after accounting for the effects of other covariates (additional details are provided in the Supplementary Information). Statistical significance was assessed by comparing results to those obtained through 250 random permutations of the covariates.

### Implications for isolation impact

We estimated the extent of shedding that would occur before and after a period of isolation by first calculating the proportion of total shedding that had occurred up to any given day, t, as F_v_(t), where F_v_ is the cumulative distribution of the best-fitting model for viral shedding, f_v_. The proportion of shedding that would have occurred prior to symptom onset and isolation (i.e., pre-symptomatic shedding) is F_v_(0), and the proportion of shedding that would still occur if isolation ended on day t_I_ is 1 – F_v_(t_I_). These estimates account for the magnitude of shedding, in addition to the duration, and are thus more informative than estimates of shedding duration alone.

The value of t_I_ represents the duration of isolation and was assigned for each individual to align with CDC recommendations for staying at home: t_I_ was set to 24 hours after fever resolution for those who experienced fever, and after 4 (or 5) days post symptom onset for those who did not experience fever [2, 3]. The ‘back translation’ property of S_ILI_ was used to identify individuals who experienced fever (i.e., attained S_ILI_ >2) and to determine the time of fever resolution (i.e., when the fitted S_ILI_ trajectory first decreased below 2 after attaining its peak). Note that the fitted S_ILI_ trajectories are a continuous representation of a discrete scoring system and so although fever is assigned a value of 3 in S_ILI_, anything greater than 2 (i.e. anything above the score assigned for cough + sore throat) is interpreted as possible fever for the purpose of this analysis. In general, S_ILI_ >2 was a faithful indicator of those who experienced fever (Supplementary Information).

Given that Ct values reflect total virus genomic material in a specimen rather than infectious shed virus (i.e., virus with onward transmission potential) we also sought to translate our estimates of viral shedding remaining following the end of isolation to estimates of infectious shedding, or transmission potential, remaining. First, we defined several candidate functions, g_v_(t), to describe the proportion of Ct-measured virus that is infectious at any given time. These included (i) a worst-case scenario in which 100% of Ct-measured virus is infectious for the entire duration of shedding; (ii) a best-case scenario in which 100% of Ct-measured virus is infectious until 4 days post symptom onset, and then 0% for the remainder; and (iii) an intermediate scenario in which the infectious percentage of Ct-measured virus decreases linearly from 0 to 7 days following symptom onset [10, 30, 37, 38]. These functions were designed for illustrative purposes and are not an exhaustive list of possible infectious virus dynamics. We then estimated the percentage of transmission potential remaining at time t_I_ as (1 – G_f_(t_I_)) x 100%, where G_f_(t) is the cumulative distribution of g_v_(t) x f_v_(t). Assuming secondary infections caused by an infectious individual are distributed in proportion to their infectious shedding distribution, the number of secondary infections generated following the end of isolation can be approximated as R_t_ x (1 – G_f_(t_I_)), where R_t_ is the effective reproduction number.

Model fitting and parameter estimation were conducted using Monolix 2021R2 and the `lixoftConnectors` and `Rsmlx` packages in R 4.0.3 (Table S1) [39–42]. Downstream analyses and plotting were conducted with the `hier.part`, `tidyverse`, `here`, `patchwork`, `ggpubr` and `scico` packages [43–48]. Further methodological details are provided in the Supplementary Information.

## Results

We identified 116 household contacts with PCR-detectable influenza virus infection and at least two Ct values <40 (Figure S2). Of these, 108 (93%) reported at least one symptom and 105 (91%) reported at least one ILI symptom (fever, cough, or sore throat) during study follow-up (Table S2). Most infections were caused by influenza A viruses (86/116; 74%), and 48 (41%) individuals were vaccinated with the current season’s influenza vaccine. Additional characteristics partitioned by age and vaccination status or influenza-like-illness (ILI) symptoms are provided in Table S3.

For individuals experiencing at least one ILI symptom (N = 105), a Weibull distribution with shape parameter (a) and magnitude parameter (h) modified by age provided the best-fit to the Ct data, whereas a Weibull distribution with scale parameter (b) modified by age and magnitude modified by vaccination status provided the best-fit to the S_ILI_ scores (Table S4, Figures S3-S5). Results were similar when fitting to the Ct and S_ANY_ (or S_UNW_) observations from those who reported any symptom (N = 108; Table S5, Figures S5-S8). These findings suggest viral shedding dynamics differ by age, and symptom score dynamics differ by age and vaccination status.

To investigate further, we tested whether age or vaccination status were associated with any trajectory summary metrics. First, we found that young children (aged <5 years) shed significantly more total virus (p < 0.0001), had higher peaks in shedding (p < 0.0001), and had longer durations of shedding (p < 0.0001) relative to other age groups (Figure 2, Figure S9). Children aged <5 years also experienced higher peaks in S_ILI_ than adults aged ≥50 years and shorter durations of S_ILI_ than adults ≥18 years (Figure S10, p < 0.01). Conversely, there were no differences in peak symptom score by age when using S_ANY_ or S_UNW_ (Figures S11-12). This was likely driven by the contribution of wheezing and shortness of breath to S_ANY_ and S_UNW_: older individuals, who were less likely to experience fever (Figure S10), were still assigned higher symptom scores if they reported these additional symptoms (Figure S11). Thus, the S_ANY_ or S_UNW_ scores may be more sensitive to symptom progression in older individuals.

**Figure 2.**
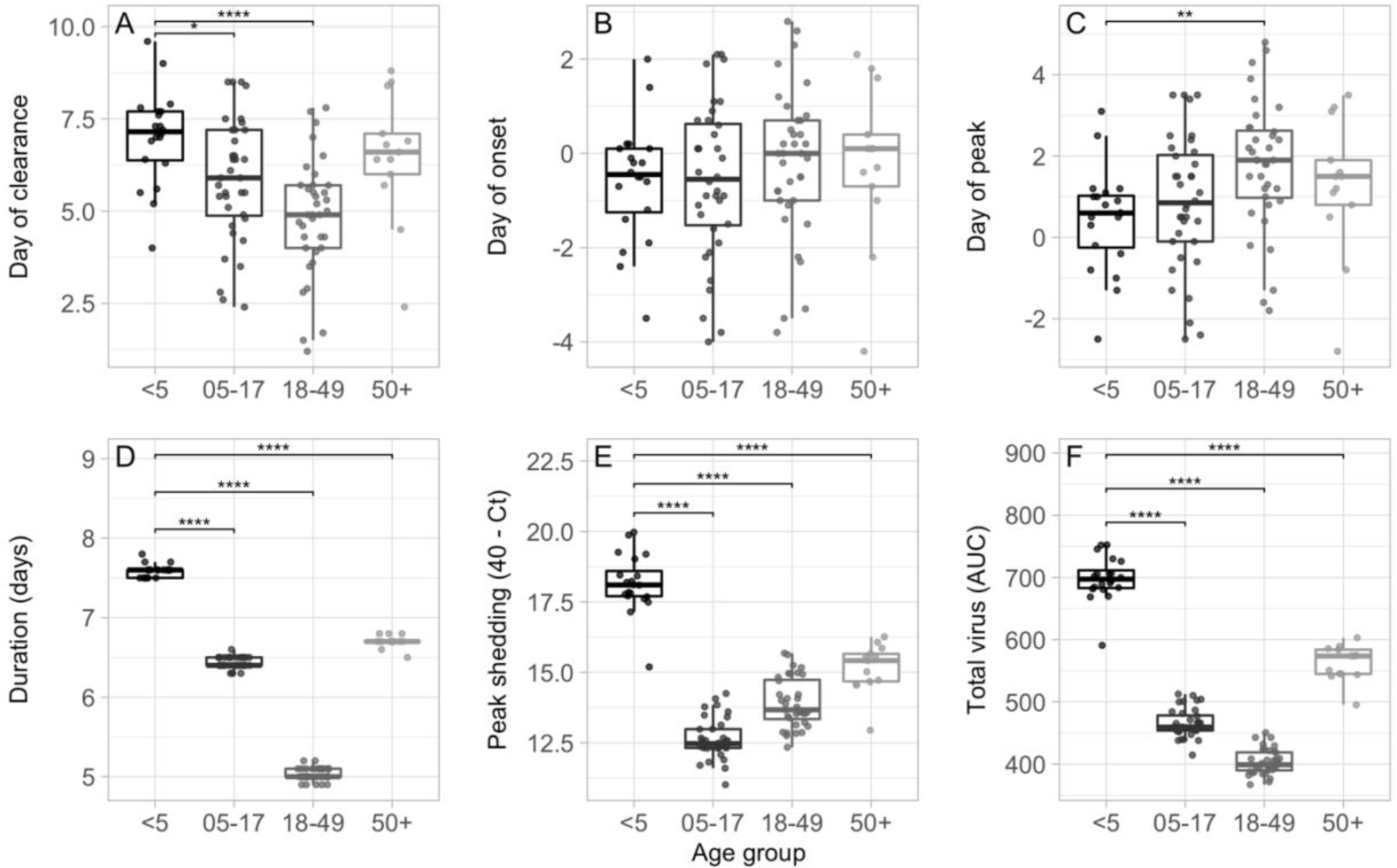
Children aged <5 years shed more virus than other age groups. Associations between age group and viral shedding summary metrics: (A) day of shedding clearance relative to ILI onset; (B) day of shedding onset relative to ILI onset; (C) day of peak shedding relative to ILI onset; (D) duration of shedding in days; (E) Peak value of shedding attained (transformed as 40 – Ct); and (F) total virus shed, as measured by the area under the fitted shedding curve. ILI represents influenza-like-illness and AUC represents the area under the curve.*p <0.05, **p <0.01, ***p <0.001, ****p <0.0001.

There were no significant associations between vaccination status and the viral shedding metrics (Figure S13). This is unlikely to be confounded by age as the hierarchical partitioning analysis also found no independent effect of vaccination status on the shedding summary metrics (Figure S14). In contrast, vaccinated individuals experienced lower total S_ILI_ scores (p = 0.02) than unvaccinated individuals (Figure 3). Hierarchical partitioning confirmed that this observation was unlikely to be confounded by age (Figure S14). Results were similar with S_UNW,_ although vaccinated individuals also experienced shorter durations in S_UNW_ symptom scores (p <0.01) (Figure S15). Conversely, we found no systematic relationship between the summary metrics and vaccination status with S_ANY_ (Figure S16).

**Figure 3.**
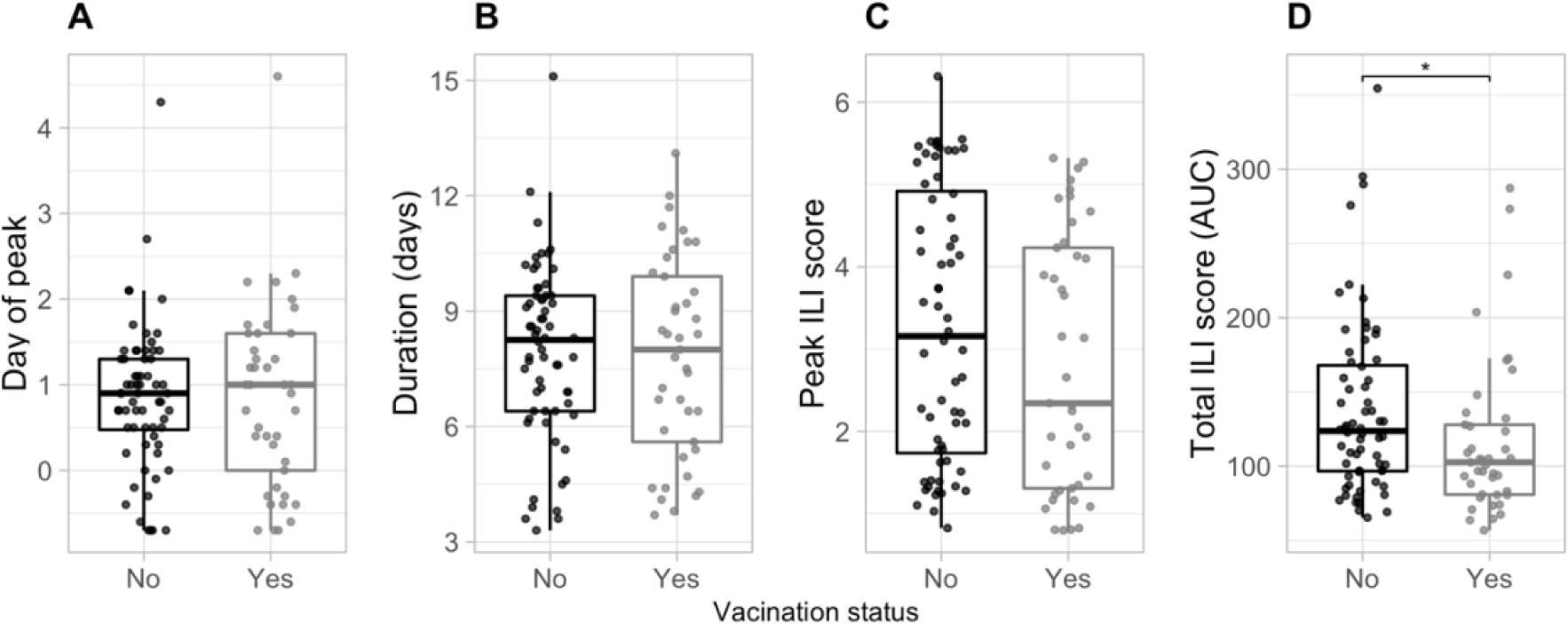
Vaccinated individuals experience lower. S_ILI_ **scores.** Associations between vaccination status and S_ILI_ summary metrics: (A) time of peak S_ILI_ score relative to days since ILI onset; (B) duration of S_ILI_ score in days; (C) peak S_ILI_ score attained; and (D) total S_ILI_ score, as measured by the area under the fitted S_ILI_ symptom curve. ILI represents influenza-like-illness and AUC represents the area under the fitted symptom score curve. *p <0.05, **p <0.01, ***p <0.001, ****p <0.0001.

Among the other covariates tested (virus type and season), we found an association between season and the time of shedding: infections from 2017-2018 experienced later peaks in shedding (p <0.01) than infections from 2018-2019 and 2019-2020 (Figure S17A). However, the hierarchical partitioning analysis suggested this association may be confounded by age (Figure S14), since children <5 were not represented in the 2017-18 data (Figure S1) and may experience earlier peaks in shedding (Figure 2). We also did not detect any associations between shedding among individuals who reported higher ILI scores (peak S_ILI_ >2) and those who reported lower scores (peak S_ILI_ ≤2) (Figure S17B).

Finally, we fit Ct data from 71 individuals with incident infection to compare shedding dynamics during asymptomatic and symptomatic infection (Figures S2 and S18). Asymptomatic individuals experienced substantially shorter durations of shedding than symptomatic individuals, in addition to lower peak and total shedding levels (Figure S19). We did not include additional covariates or formally test these results due to the sample size (8 asymptomatic infections and 63 symptomatic infections).

### Implications for isolation impact

Any shedding that occurs prior to symptom onset (‘pre-symptomatic shedding’) will reduce the effectiveness of isolation measures. In this study, we estimated that most individuals (60%; 63/105) shed less than 10% of their total virus before the onset of an ILI symptom, although 15% (16/105) shed more than 50% before onset (Figure 4A). Individuals in the latter group had no characteristic in common with respect to age, vaccination status, type of infecting virus, or season of infection (Table S6). Across all individuals there was a nonsignificant reduction in average pre-symptomatic shedding among adults ≥18 years compared to children <18 years (p = 0.2; Figure 4B), and no relationship with vaccination status (Figure 4C).

**Figure 4.**
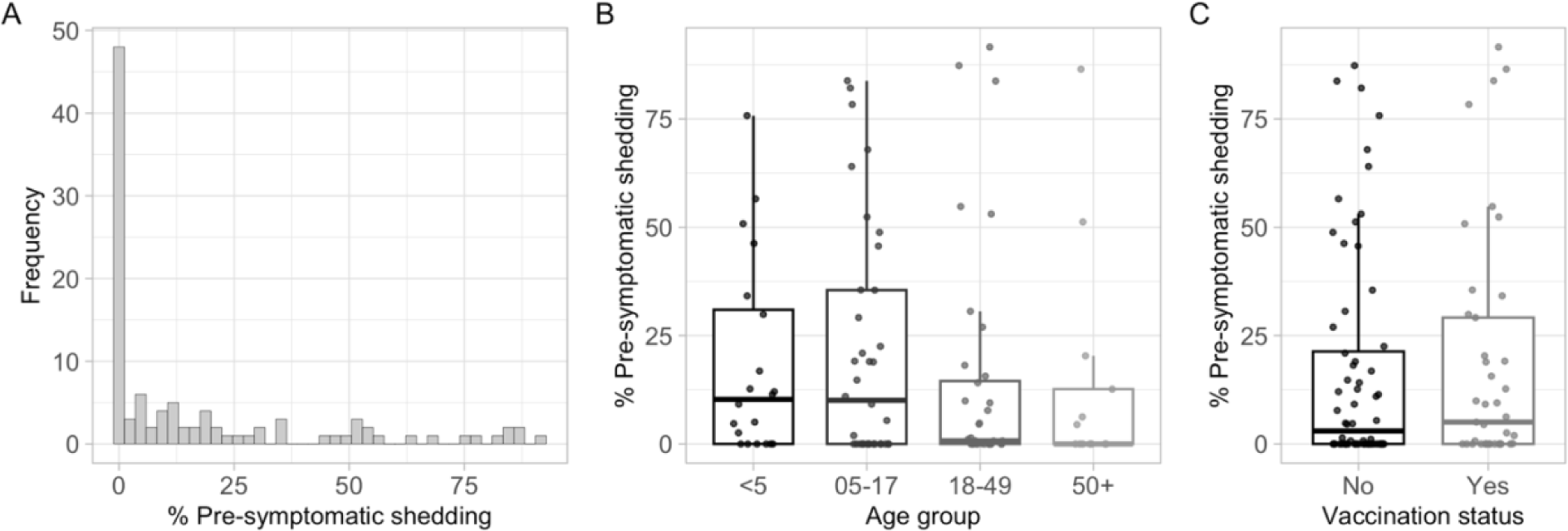
Pre-symptomatic shedding is heterogeneous. (A) Distribution of pre-symptomatic shedding across all individuals with ILI symptoms (N = 105). (B) Association between pre-symptomatic shedding and age group. (C) Association between pre-symptomatic shedding and vaccination status. Pre-symptomatic shedding was estimated as the percentage of total virus shedding that occurred prior to ILI symptom onset. ILI stands for influenza-like-illness. *p <0.05, **p <0.01, ***p <0.001, ****p <0.0001.

In addition to pre-symptomatic shedding, shedding may also persist after isolation has ended. Current recommendations advise symptomatic individuals infected with influenza viruses to stay at home and avoid contact with others until 24 hours after fever resolution, or 4-5 days following symptom onset if they do not experience fever [2, 3]. We henceforth refer to the former as a ‘fever-based’ strategy and the latter as a ‘duration-based’ strategy. For those following the fever-based strategy (i.e., those who experienced fever), we calculated the percentage of shedding remaining one day post fever resolution (Figures 5A-B). For those who did not experience fever and instead followed the duration-based strategy, we calculated the percentage of shedding remaining after four days post ILI symptom onset (i.e., on day five post ILI symptom onset; Figure 5C). We considered shedding remaining after five days post ILI symptom onset in sensitivity analyses (Figure S20). We then combined estimates across all individuals to understand post-isolation shedding distributions at the population-level, if isolation practices aligned with current recommendations for staying at home. In this study, the median percentage of shedding remaining after isolation was 1-29% across age and vaccination groups, and there was substantial inter-individual variation in shedding remaining and in isolation duration (Figures 5D-E). Notably, adults ≥50 years were less likely to experience fever and thus more likely to isolate for a fixed duration of four days. We also identified eight individuals with fever would have isolated for over six days. If all symptomatic participants instead followed the four-day duration-based strategy (i.e., everyone isolated for four days following ILI symptom onset), average estimates of shedding remaining were equal to, or lower than, the combination of fever-based and duration-based strategies, and inter-individual variation was greatly reduced (Figure 5F).

**Figure 5.**
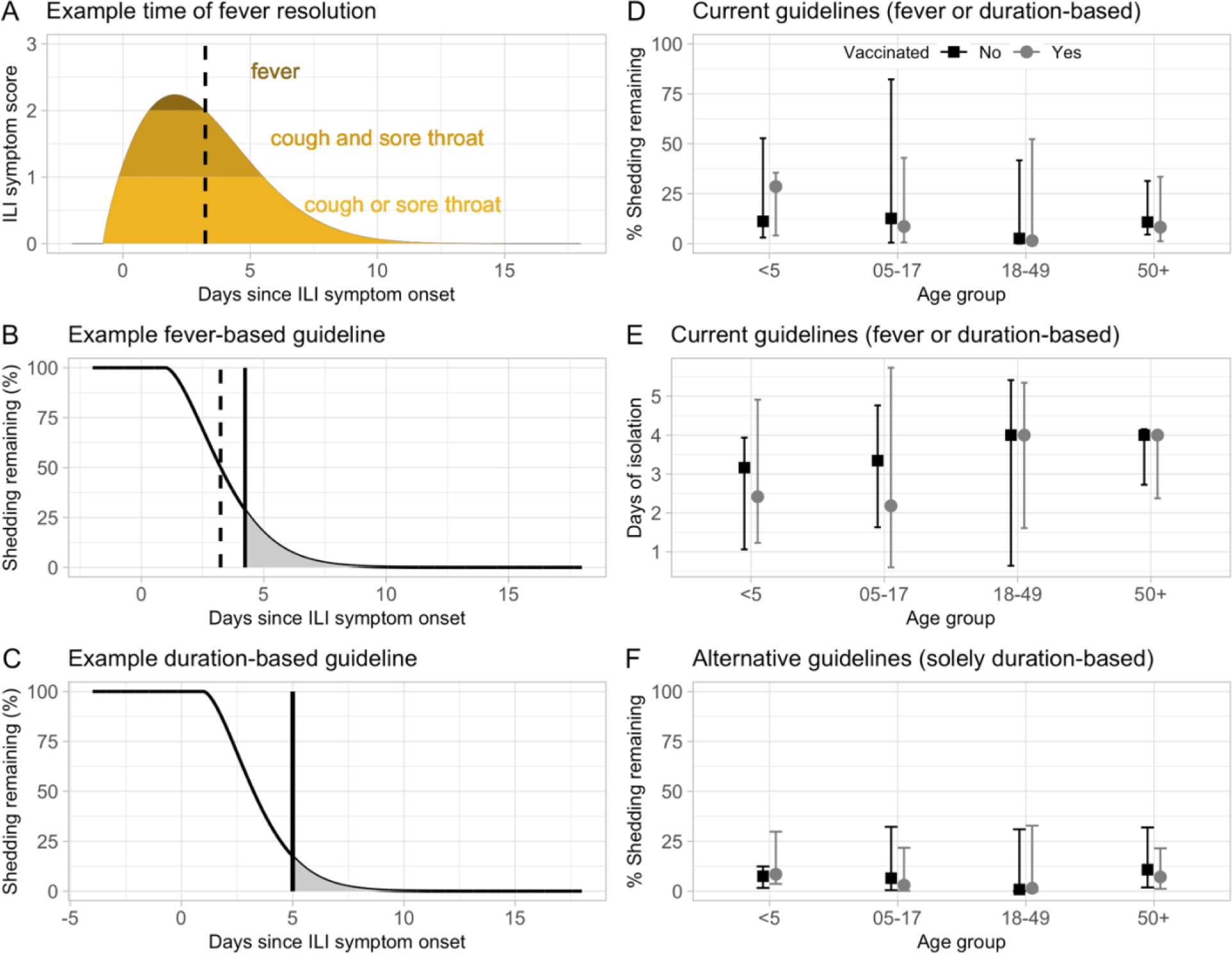
Duration-based isolation strategies are effective in reducing shedding remaining after isolation from symptomatic individuals. (A-B) Example ILI symptom (A) and shedding remaining (B) trajectories from a child aged 5-17 years following a fever-based isolation strategy. The dashed vertical line represents the time of fever resolution, and the solid vertical line represents the end of recommended isolation. (C) Corresponding shedding remaining trajectory for the same child following a duration-based isolation strategy. (D-E) Estimates of shedding remaining (D) and duration of isolation in days (E) assuming isolation practices align with current recommendations for staying at home (i.e., a mixture of fever-based and duration-based strategies). (F) Estimates of shedding remaining assuming all individuals follow a four day duration-based isolation strategy, including those who experience fever. Estimates in (E, F) are stratified by age and vaccination status. Points represent the median within each stratification and error bars are the 90^th^ percentiles.

Ct trajectories reflect the dynamics of total viral RNA in a sample rather than infectious virus (i.e., virus with onward transmission potential). We therefore explored how estimates of shedding remaining could be translated to estimates of infectious shedding, or transmission potential, remaining by defining three candidate functions, g_v_(t), to describe the proportion of Ct-measured virus that is infectious at any given time: (i) a worst-case scenario in which all shed virus was assumed infectious; (ii) a best-case scenario in which all shed virus was infectious until four days post ILI symptom onset and 0% was infectious following that time; and an intermediate scenario in which the infectiousness of shed virus decayed linearly from 0-7 days post ILI symptom onset (Figures 6A-B) [10, 30, 37, 38]. Unsurprisingly, estimates of infectious shedding remaining depend on the assumed infectiousness function, g_v_(t): for an example individual ending isolation after four days post ILI symptom onset, the percentage of infectious virus remaining was 0%, 5% and 17% in the best, intermediate, and worst-case scenarios, respectively (Figure 6C). Translating these estimates to the potential number of secondary infections generated per infected individual following the end of a four-day duration-based isolation strategy (assuming R_t_ = 1.2 [49]) resulted in projections ranging from 0–0.6 across participants (Figure 6D). Given the strong assumptions underlying each infectiousness function, these results are presented for illustration purposes only.

**Figure 6.**
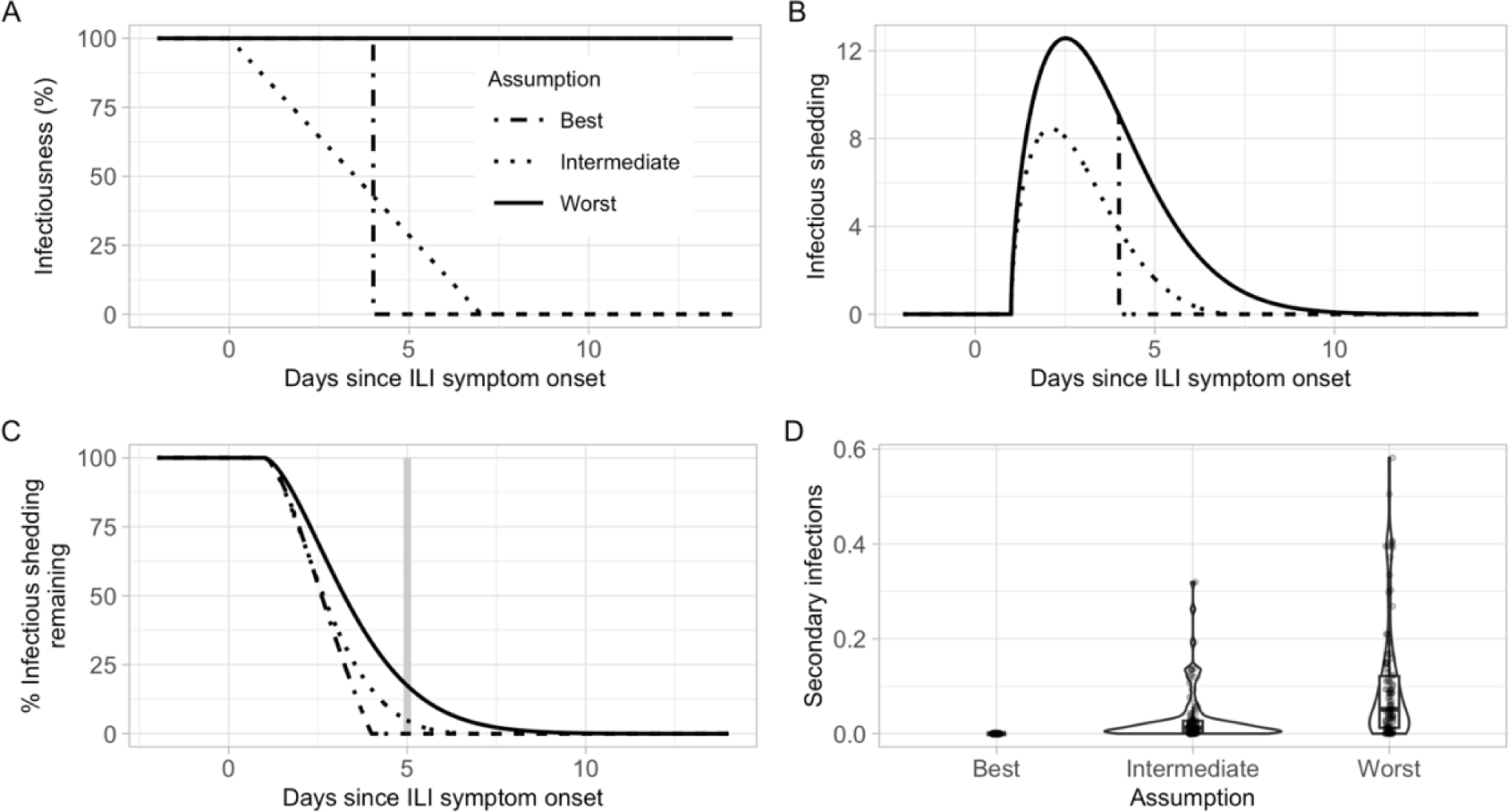
Framework for translating viral shedding to estimates of onward transmission potential. (A) Example functions for the percentage of shed virus that is infectious at any time, g_v_(t). (B) Example application of functions for g_v_(t) shown in (A) to the fitted Ct trajectory of the individual shown in Figure 5B. The worst-case scenario reflects the original fitted Ct trajectory. (C) Corresponding estimates of the percentage of infectious shedding remaining for the same individual and the same functions for g_v_(t) shown in (A). The grey vertical line indicates the end of a four day duration-based isolation strategy. (D) Estimates of transmission potential remaining across all symptomatic individuals following the end of a four day duration-based isolation strategy, expressed as the number of secondary infections caused by each individual when R_t_ = 1.2.

## Discussion

The effectiveness of isolation strategies for individuals with symptomatic influenza virus infection depends in part on the dynamics of virus shedding in relation to symptom timing and severity. We fit nonlinear mixed-effects models to data from an influenza household transmission study and identified differences in shedding by age and symptom scores by age and vaccination status. We also found relatively low levels of pre-symptomatic, asymptomatic, and post-isolation shedding, which could have important implications for the effectiveness of influenza control through isolation.

Young children are thought to play a central role in influenza transmission within households due to increased susceptibility and infectivity [6, 16, 50]. Here, we found that young children aged <5 years shed more virus than all other age groups and experienced higher peaks and longer durations of shedding. A previous study also found children (aged ≤18 years) experienced longer durations of shedding than adults, although there was no difference in shedding levels at symptom onset [30]. The difference between the latter result and our identified association with peak shedding levels may be influenced by how age groups were partitioned. The significantly higher peak shedding in children aged <5 years found in this study would have likely been obscured if young children and adolescents (aged 5-17 years) had been analyzed together as previously reported [30]. In addition to differences in shedding dynamics, we identified age-related differences in symptom duration and severity. The former increased with age and has been reported previously [7]. The latter was influenced by the chosen symptom score: S_ILI_ scores tended to decrease with age whereas S_ANY_ and S_UNW_ scores did not. This is likely due to the inclusion of wheezing and shortness of breath in S_ANY_ and S_UNW_, which may offer a more sensitive description of symptom burden in older adults for whom fever is less common.

In contrast to age, we found no difference in shedding dynamics by vaccination status. This suggests vaccination did not reduce transmission potential in participants with breakthrough infections. Our finding aligns with results from a previous household transmission study [37] and can help inform mathematical modeling studies which must typically make assumptions regarding the infectiousness of breakthrough infections in vaccinated individuals relative to unvaccinated individuals. Despite similar shedding profiles, vaccinated infected participants did experience lower total S_ILI_ scores than their unvaccinated counterparts. This effect was likely driven by a reduction in fever, given that the association was present with S_UNW_ (which also includes an independent term for fever) but not with S_ANY_ (in which wheezing or shortness of breath can compensate for the absence of fever). Our results are consistent with a meta-analysis that found influenza-infected vaccinated individuals were significantly less likely to develop fever than infected unvaccinated individuals, but equally likely to develop other symptoms including cough, headache, sore throat, wheezing, fatigue, and nasal congestion [51]. More generally, our findings highlight the importance of considering multiple scoring metrics when analyzing individual symptom dynamics.

In addition to identifying individual differences in shedding and symptom dynamics, we explored the potential impact of these dynamics on the effectiveness of influenza isolation measures by estimating levels of pre-symptomatic and post-isolation shedding from symptomatic individuals, and by comparing shedding dynamics of symptomatic individuals with those of asymptomatic individuals (who would not be reached by isolation recommendations). We found pre-symptomatic shedding levels were relatively low for most participants, consistent with previous work [52]. However, 15% experienced more than 50% of their shedding prior to symptom onset, suggesting isolation could miss a substantial proportion of transmission in a small fraction of individuals. With respect to post-isolation shedding, although longer durations of isolation will generally favor greater reductions in shedding, this effect must be balanced with the financial and emotional burdens associated with adhering to such measures for prolonged periods [53]. Notably, we found that isolation of all symptomatic persons for four days post ILI symptom onset, regardless of the presence of fever, may be as effective in reducing shedding compared with a strategy informed by current recommendations for staying at home (which had different recommendations for people experiencing fever and would have required some participants to isolate for six days or more). Finally, we found that asymptomatic infections were rare and shed substantially less virus, for shorter periods of time, than symptomatic infections. Although the sample size was small, these findings support previous analyses [13, 52] and suggest that transmission from symptomatic individuals could comprise the majority of seasonal influenza transmission. There may also be implications for mathematical modeling studies which often assume symptomatic and asymptomatic infections are equally transmissible [54]. Overall, our results suggest prompt isolation of symptomatic individuals could be an effective measure for influenza control due to generally low levels of pre-symptomatic transmission, virus shedding post-isolation, and shedding from asymptomatic individuals.

There are several limitations to this study. First, shedding trajectories were fit to Ct values which detect total viral RNA in a sample rather than infectious shed virus. Quantifying the latter would require additional data, such as viral culture, that were not available for this study. Given that influenza viral culture measures decline more rapidly than total shed virus [37], our estimates of pre-symptomatic and post-isolation shedding are likely upper bounds on infectious virus shedding. Although these can still be useful for planning purposes [23], modeling dynamics of infectious virus, and assessing whether these differ by age and/or other covariates, is a critical avenue for future work. Similarly, when estimating the number of secondary infections that occur after isolation, we assumed these were distributed in proportion to an individual’s estimated infectious shedding distribution. Although this may be largely consistent with transmission in community settings, where isolation guidance is most applicable, it does not account for typically higher rates of exposure and susceptible depletion that can occur within households.

Second, we used symptom diaries to quantify symptom onset and severity, which may include reports of symptoms caused by factors other than influenza virus infection (henceforth referred to as non-specific symptoms). For example, individuals may be more likely to report fatigue and runny nose which have many other causes. Reporting of non-specific symptoms may contribute to the high frequency of symptomatic infection identified in this study compared to others [55–57]. In addition, our estimates of pre-symptomatic shedding (which depend on the time of reported symptom onset) may be underestimated if non-specific symptoms were reported before influenza-specific symptoms. To mitigate these impacts, we used definitions that closely track influenza related symptoms (for example, ILI), and excluded data from individuals who reported symptoms on three or more days with a corresponding negative influenza test, before their first positive test. However, our symptom score trajectories may still be biased towards earlier and/or higher values.

Finally, we used a phenomenological approach to fit the functional forms of the shedding and symptom score data, rather than developing a mechanistic model to explain underlying virus replication and cell infection dynamics [58–61]. This choice was motivated by data availability: fitting an appropriate and identifiable mechanistic model for influenza virus infection requires information on viral load and various immune cell populations that were not captured in this study. Although our approach cannot estimate within-host parameters such as viral growth rates and infected cell lifetimes, we are still able to capture shedding and symptoms score trajectories and explore differences in their relative timing and magnitude.

In this work we have illustrated how phenomenological modeling of data routinely collected during household transmission studies can elucidate virus shedding and symptom dynamics within individuals and identify differences by age and vaccination status. Our framework can also be used to estimate the impact of isolation in reducing influenza virus transmission, and thus inform effective strategies for influenza isolation.

## Data Availability

All data produced in the present study are available upon reasonable request to the authors.

## Acknowledgements

The authors would like to thank Alicia Fry, Scott Blackwell, Emma Pedigree-Cannon, Holly Morse, Megan Eluhu, Emily Jookar, Christina Khouri, Robert Lyons, Carleigh Frazier, Janika Raynes, Preston Gibson, Karen Malone, Sarah Davis, Olivia Doak, Judy King, and the study participants.

