## Supplementary Information for "Influenza virus shedding and symptoms: Dynamics and implications from a multi-season household transmission study"

### Supplementary materials

#### Supplementary text

##### Model fitting in Monolix

We fit our models to the data using a nonlinear mixed effects (NLME) framework in Monolix 2021R2. Monolix is used extensively in the fields of within-host modeling and pharmokinetics / pharmacodynamics and is available at https://lixoft.com/products/monolix/. We fit $\hat{V}$(t) to the Ct observations, treating negative Ct tests as censored observations, and fit $W(t)$ to the symptom score data. We assumed normal distribution of both variables with constant error terms and ensured model residuals were normally distributed using the Shapiro-Wilk test. Each parameter was allowed to vary across individuals by including both a fixed and random effect. We assumed $a, b$ and $h$ were lognormally distributed to ensure positivity, and $d$ was normally distributed to allow positive or negative shifts in time. We found no strong evidence for correlations between parameters during initial model fitting and so assumed all parameters were independent in subsequent fitting. We explored models that controlled for candidate covariates (age group, vaccination status, virus type or season) with respect to one or more parameters and evaluated the importance of each covariate-parameter relationship using ANOVA. Those relationships with p-values < 0.01 were kept in the final model. We compared models assuming different distributions for $f_v$ and $f_w$ (Weibull, Gamma, or Lognormal) using Akaike Information Criterion (AIC), where AIC $= 2k - 2lnL$, $k$ is the number of estimated parameters and $lnL$ is the maximum log likelihood.

##### Calculating trajectory summary metrics

For each fitted Ct trajectory ($\hat{V}(t)$) we estimated the onset (clearance) of shedding as the first (last) time at which $\hat{V}(t) \geq 1$. The duration of shedding was the time between these two estimates. Since these estimates vary based on the choice of threshold (here equal to 1), they are most useful in making relative comparisons between covariate groups, rather than determining absolute values.

Similarly, for each fitted symptom score trajectory ($W(t)$), the time of symptom clearance was estimated as the last time at which $W(t) \geq 0.3$. The threshold of 0.3 was used as the data are not continuous, and a score of 0.3 equals the reporting of one symptom (out of runny nose, nasal congestion, or fatigue) for $S_{ANY}$. We do not need to calculate an onset time for the symptom score trajectories as they are already estimated relative to time since symptom onset.

#### Hierarchical partitioning

##### Intuition

To distinguish the covariates with the greatest independent influence on trajectory dynamics from those acting primarily through collinearity with other variables, we performed hierarchical partitioning on the summary metrics. For each covariate, hierarchical partitioning considers all possible nested models within the full multivariate regression model derived from that covariate, then assesses the average increase in goodness-of-fit ($\Delta$GoF) achieved by including the covariate in each model. For example, to assess the influence of covariate A in a multivariate linear regression including covariates A, B, and C, one would consider the nested hierarchies (A, AB, ABC) and (A, AC, ABC), where AB indicates the sub-model with A and B as covariates, and so on. One would then calculate the $\Delta$GoF from including A in each model and take the average. This approach ensures the sum of the average influences of each covariate is equal to the total $\Delta$GoF between the full and null models. Thus, the average influences form a partition of the combined explanatory power of all covariates, and the percentage importance of each covariate is its average $\Delta$GoF over the total $\Delta$GoF multiplied by 100. One advantage of this approach is that the percentage importance returned for each covariate reflects its independent influence on the dependent variable, having averaged out effects of collinearity with other covariates. Furthermore, by considering all possible models within each hierarchy, the approach is not sensitive to the order in which models are assessed, as can be the case with other algorithms such as stepwise elimination.

##### Example

Here we outline the hierarchical partitioning approach for a regression model with three independent variables, taken from Chevan & Sutherland (1991). Consider a multivariate linear regression model with dependent variable Y and independent variables A, B and C. Let $\mathrm{M_{ij}}$ denote the model including $i$ and $j$, for $i, j \in$ (A, B, C), and let $\mathrm{X_{ij}}$ denote the corresponding goodness-of-fit value (GoF). Similarly, let $\mathrm{M_{ABC}}$ denote the full regression model and $\mathrm{M_0}$ the null model, and $\mathrm{X_{ABC}}$, $\mathrm{X_0}$ the corresponding GoFs, respectively.

First consider all the nested hierarchies within the full regression model, $\mathrm{M_{ABC}}$, and their corresponding GoFs. Ignoring the null model, we can list these hierarchies as follows:

| Hierarchy 1 ($\mathrm{H_{A1}}$) | Hierarchy 2 ($\mathrm{H_{A2}}$) | Hierarchy 3 ($\mathrm{H_{B1}}$) | Hierarchy 4 ($\mathrm{H_{B2}}$) | Hierarchy 5 ($\mathrm{H_{C1}}$) | Hierarchy 6 ($\mathrm{H_{C2}}$) |
|---|---|---|---|---|---|
| $\mathrm{X_A}$ | $\mathrm{X_A}$ | $\mathrm{X_B}$ | $\mathrm{X_B}$ | $\mathrm{X_C}$ | $\mathrm{X_C}$ |
| $\mathrm{X_{AB}}$ | $\mathrm{X_{AC}}$ | $\mathrm{X_{AB}}$ | $\mathrm{X_{BC}}$ | $\mathrm{X_{AC}}$ | $\mathrm{X_{BC}}$ |
| $\mathrm{X_{ABC}}$ | $\mathrm{X_{ABC}}$ | $\mathrm{X_{ABC}}$ | $\mathrm{X_{ABC}}$ | $\mathrm{X_{ABC}}$ | $\mathrm{X_{ABC}}$ |

We can see that hierarchies $\mathrm{H_{A1}}$ and $\mathrm{H_{A2}}$ represent subsets in which all models include the variable A, $\mathrm{H_{B1}}$ and $\mathrm{H_{B2}}$ are subsets in which all models include B, and $\mathrm{H_{C1}}$ and $\mathrm{H_{C2}}$ are subsets in which all models include C. Thus we refer to A as the principle variable for hierarchies $\mathrm{H_{A1}}$ and $\mathrm{H_{A2}}$, B the principal variable for $\mathrm{H_{B1}}$ and $\mathrm{H_{B2}}$, and so on.

For each model, then consider the increase in GoF ($\Delta$GoF) obtained by including the principle variable. These differences can be expressed as

| $H_{A1}$ | $H_{A2}$ | $H_{B1}$ | $H_{B2}$ | $H_{C1}$ | $H_{C2}$ |
|---|---|---|---|---|---|
| $X_A$ - $X_0$ | $X_A$ - $X_0$ | $X_B$ - $X_0$ | $X_B$ - $X_0$ | $X_C$ - $X_0$ | $X_C$ - $X_0$ |
| $X_{AB}$ - $X_B$ | $X_{AC}$ - $X_C$ | $X_{AB}$ - $X_A$ | $X_{BC}$ - $X_C$ | $X_{AC}$ - $X_A$ | $X_{BC}$ - $X_B$ |
| $X_{ABC}$ - $X_{BC}$ | $X_{ABC}$ - $X_{BC}$ | $X_{ABC}$ - $X_{AC}$ | $X_{ABC}$ - $X_{AC}$ | $X_{ABC}$ - $X_{AB}$ | $X_{ABC}$ - $X_{AB}$ |

Let $S_{A1}$ denote the sum of the ΔGoFs for hierarchy $H_{A1}$, $S_{A2}$ the sum of the ΔGoFs for hierarchy $H_{A2}$, and so on. Then

$$S_{\mathrm{A1}} = (X_{\mathrm{A}} - X_0) + (X_{\mathrm{AB}} - X_{\mathrm{B}}) + (X_{\mathrm{ABC}} - X_{\mathrm{BC}})$$
$$S_{\mathrm{A2}} = (X_{\mathrm{A}} - X_0) + (X_{\mathrm{AC}} - X_{\mathrm{C}}) + (X_{\mathrm{ABC}} - X_{\mathrm{BC}}).$$

Similar expressions can be written for $S_{B1}$, $S_{B2}$, $S_{C1}$ and $S_{C2}$.

The average ΔGoF for all nested hierarchies in which A is the principle variable, $D_A$, is then given by

$$\begin{aligned} D_{\mathrm{A}} &= \frac{S_{\mathrm{A1}} + S_{\mathrm{A2}}}{6} \\ &= \frac{(X_{\mathrm{A}} - X_0) + (X_{\mathrm{AB}} - X_{\mathrm{B}}) + (X_{\mathrm{ABC}} - X_{\mathrm{BC}}) + (X_{\mathrm{A}} - X_0) + (X_{\mathrm{AC}} - X_{\mathrm{C}}) + (X_{\mathrm{ABC}} - X_{\mathrm{BC}})}{6} \\ &= \frac{2X_{\mathrm{ABC}} + X_{\mathrm{AB}} + X_{\mathrm{AC}} - 2X_{\mathrm{BC}} - X_{\mathrm{B}} - X_{\mathrm{C}} + 2X_{\mathrm{A}} - 2X_0}{6}. \end{aligned}$$

It follows that if we sum the average ΔGoFs across all variable hierarchies we get

$$\begin{aligned} D_{\mathrm{A}} + D_{\mathrm{B}} + D_{\mathrm{C}} &= \frac{S_{\mathrm{A1}} + S_{\mathrm{A2}} + S_{\mathrm{B1}} + S_{\mathrm{B2}} + S_{\mathrm{C1}} + S_{\mathrm{C2}}}{6} \\ &= \frac{6X_{\mathrm{ABC}} - 6X_0}{6} \\ &= X_{\mathrm{ABC}} - X_0. \end{aligned}$$

So the sum of the average ΔGoFs for each variable is equal to the total ΔGoF between the full and null models. Thus the average ΔGoFs ($D_A$, $D_B$ and $D_C$) partition the combined explanatory power of the full model among each of the independent variables, A, B and C.

#### Identifying fever from $S_{ILI}$

The fitted $S_{ILI}$ trajectories are a continuous representation of a discrete scoring system and so although fever is assigned a value of 3 in $S_{ILI}$, anything greater than 2 (i.e. anything above the score assigned for cough + sore throat) is interpreted as possible fever for the purpose in our analysis. However, instances where the fitted $S_{ILI}$ trajectory for an individual who did *not* report fever attained a value greater than 2 were relatively rare (6/68 fitted trajectories; 9%). Similarly, just 1/63 (2%) individuals who reported fever had an $S_{ILI}$ trajectory that did *not* attain a value greater than 2. Thus the $S_{ILI} > 2$ threshold is a faithful means of identifying occurrences of fever from fitted ILI symptom score data.

#### Supplementary tables

**Table S1 – Initial conditions for nonlinear mixed-effects model fitting in Monolix.** All other parameters were set to default values.

| Parameter | Model | Fixed effect initial value |
|---|---|---|
| Shape | Ct shedding relative to ILI onset | 2 |
| Scale | Ct shedding relative to ILI onset | 1.5 |
| Magnitude | Ct shedding relative to ILI onset | 50 |
| Shift | Ct shedding relative to ILI onset | 0 |
| Shape | Ct shedding relative to alternative onset | 2 |
| Scale | Ct shedding relative to alternative onset | 1.5 |
| Magnitude | Ct shedding relative to alternative onset | 50 |
| Shift | Ct shedding relative to alternative onset | 0 |
| Shape | Ct shedding relative to first positive test (for asymptomatic infections) | 2 |
| Scale | Ct shedding relative to first positive test (for asymptomatic infections) | 1 |
| Magnitude | Ct shedding relative to first positive test (for asymptomatic infections) | 4 |
| Shift | Ct shedding relative to first positive test (for asymptomatic infections) | 0 |
| Shape | ILI symptom score | 1.5 |
| Scale | ILI symptom score | 3 |
| Magnitude | ILI symptom score | 10 |
| Shift | ILI symptom score | 0 |
| Shape | Alternative symptom score | 1.5 |
| Scale | Alternative symptom score | 3 |
| Magnitude | Alternative symptom score | 15 |
| Shift | Alternative symptom score | 0 |
| Shape | Unweighted symptom score | 1.5 |
| Scale | Unweighted symptom score | 3 |
| Magnitude | Unweighted symptom score | 15 |
| Shift | Unweighted symptom score | 0 |

**Table S2 – Characteristics of household contacts infected with influenza viruses included in analysis (N = 116).** Concurrent breakdowns by age and vaccination status or influenza-like-illness (ILI) symptoms are provided in Table S3.

| Covariate | Number (%) |
|---|---|
| Age (years) | |
| <5 | 22 (19) |
| 5-17 | 41 (35) |
| 18-49 | 39 (34) |
| ≥50 | 14 (12) |
| Current season influenza vaccination status | |
| Not vaccinated | 68 (59) |
| Vaccinated | 48 (41) |
| Type | |
| Influenza A | 86 (74) |
| Influenza B | 30 (26) |
| Symptoms | |
| Reported any ILI symptom | 105 (91) |
| Reported any other symptom without ILI | 3 (2) |
| Reported no symptoms (asymptomatic) | 8 (7) |

**Table S3 – Individual breakdown by age and influenza vaccination status or ILI symptom reporting (N = 116).**

| Age (years) | Current season vaccination status | Number (%) | Reported any ILI symptom | Number (%) |
|---|---|---|---|---|
| <5 | No | 11 (9) | No | 2 (2) |
| <5 | Yes | 11 (9) | Yes | 20 (17) |
| 5-17 | No | 27 (23) | No | 5 (4) |
| 5-17 | Yes | 14 (12) | Yes | 36 (31) |
| 18-49 | No | 25 (22) | No | 3 (3) |
| 18-49 | Yes | 14 (12) | Yes | 36 (31) |
| 50+ | No | 5 (4) | No | 1 (1) |
| 50+ | Yes | 9 (8) | Yes | 13 (11) |

**Table S4 – AIC comparison of model fits to 105 trajectories relative to ILI symptom onset.** The difference in AIC, ΔAIC, for model i is calculated as the difference in AIC value between model i and the best-fitting model with the lowest AIC. Thus, ΔAIC = 0 for the best-fitting model. A difference greater than 2 between two models indicates greater statistical support for the model with lower AIC.

| Distribution | ΔAIC, Ct model* | ΔAIC, $S_{ILI}$ model** |
|---|---|---|
| Weibull | 0 | 0 |
| Gamma | 71.1 | 17.1 |
| Lognormal | 72.8 | 37.5 |

* Shape and magnitude parameters modified by age

** Scale parameter modified by age and magnitude parameter modified by vaccination status

**Table S5 – AIC comparison of model fits to 108 trajectories relative to any symptom onset.** The difference in AIC, ΔAIC, for model i is calculated as the difference in AIC value between model i and the best-fitting model with the lowest AIC. Thus, ΔAIC = 0 for the best-fitting model. A difference > 2 between two models indicates greater statistical support for the model with lower AIC.

| Distribution | ΔAIC, Ct model* | ΔAIC, $S_{ANY}$ model** | ΔAIC, $S_{UNW}$ model** |
|---|---|---|---|
| Weibull | 0 | 0 | 0 |
| Gamma | 2.7 | 17.6 | 15.5 |
| Lognormal | 4.4 | 22.8 | 25.2 |

* Shape and magnitude parameters modified by age

** Scale parameter modified by age and magnitude parameter modified by vaccination status

**Table S6 – Characteristics of participants with more than 50% estimated pre-symptomatic shedding (N = 16).**

| Age group | Vaccination status | Season | Virus |
|---|---|---|---|
| <5 | Yes | 2019/20 | Infuenza B |
| <5 | No | 2019/20 | Infuenza A |
| <5 | No | 2019/20 | Infuenza A |
| 5-17 | Yes | 2017/18 | Influenza A |
| 5-17 | No | 2018/19 | Influenza A |
| 5-17 | No | 2018/19 | Influenza A |
| 5-17 | Yes | 2018/19 | Influenza A |
| 5-17 | Yes | 2018/19 | Influenza A |
| 5-17 | No | 2018/19 | Influenza A |
| 18-49 | Yes | 2018/19 | Influenza A |
| 18-49 | Yes | 2018/19 | Influenza A |
| 18-49 | No | 2019/20 | Influenza A |
| 18-49 | No | 2019/20 | Influenza B |
| 18-49 | No | 2019/20 | Influenza B |
| ≥50 | Yes | 2017/18 | Influenza A |
| ≥50 | No | 2018/19 | Influenza A |

### Supplementary figures

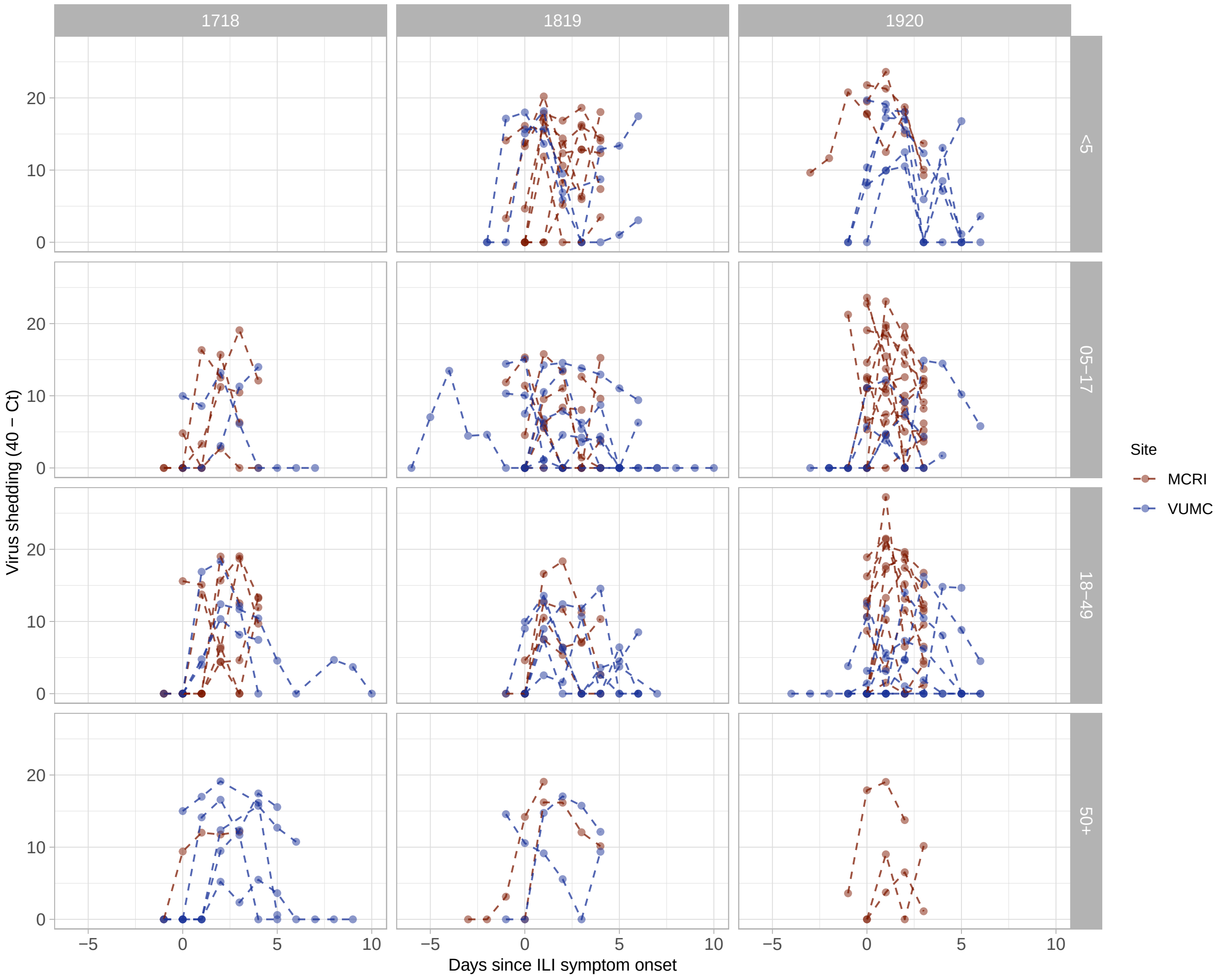


**Figure S1 – Ct values by site, age group and season.** Columns are different seasons (left to right: 2017-2018, 2018-2019, 2019-2020) and rows are different age groups (top to bottom: less than 5 years, 5-17 years, 18-49 years, and 50 years and older). MCRI stands for Marshfield Clinical Research Institute and VUMC stands for Vanderbilt University Medical Center.

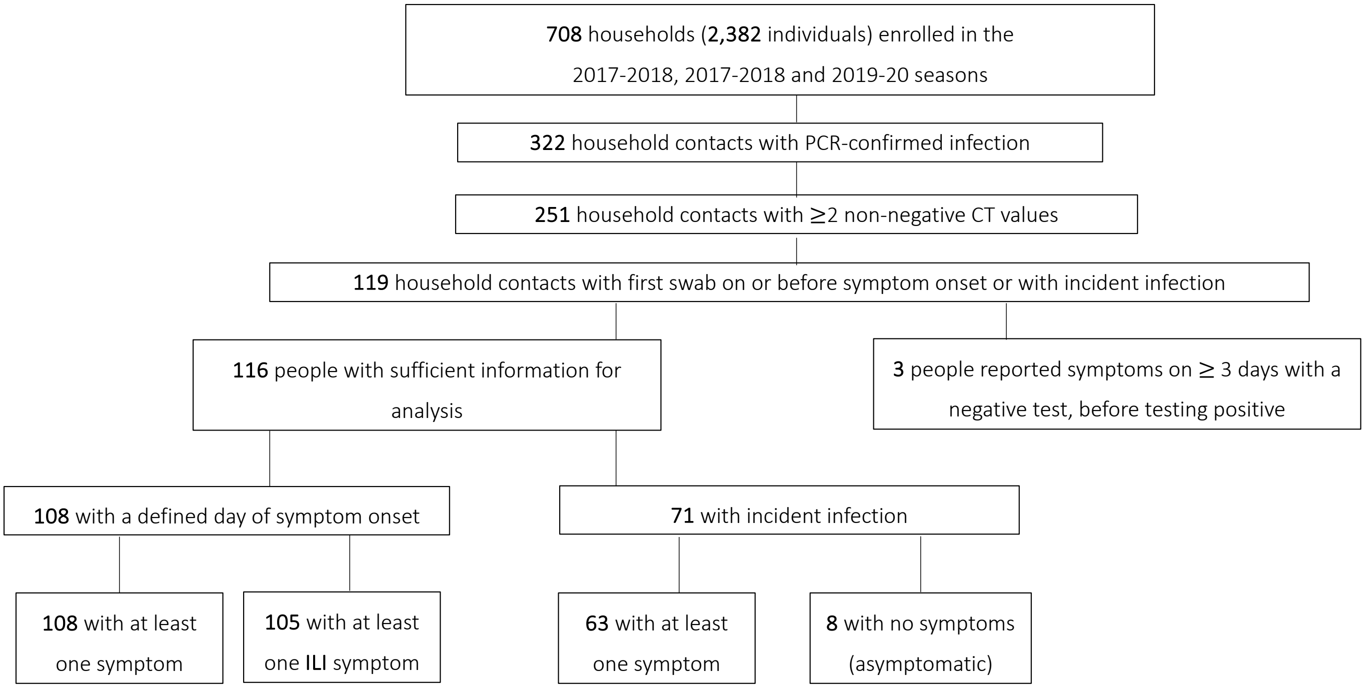


**Figure S2 – Inclusion and exclusion of individuals.** ILI stands for influenza-like-illness.

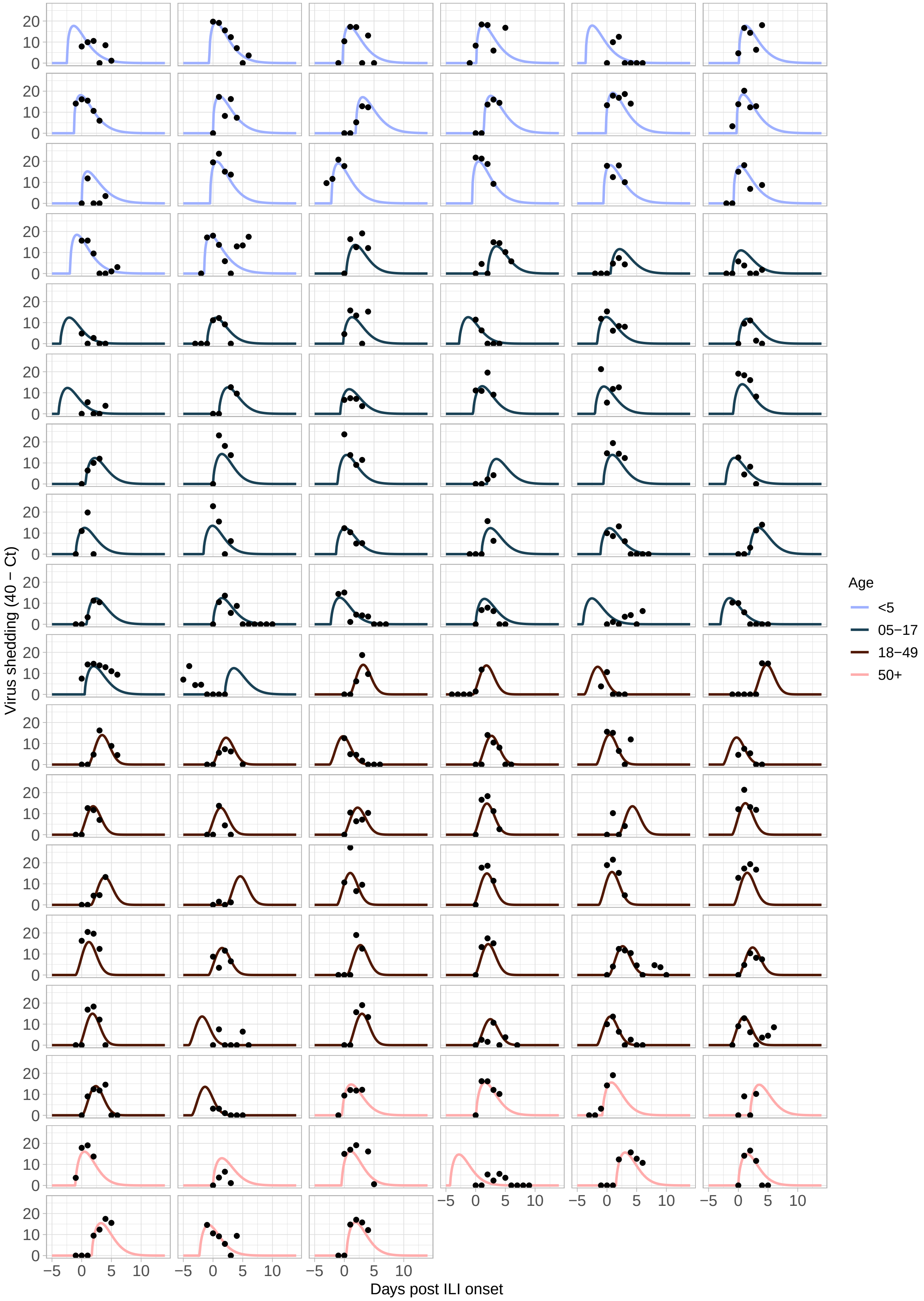


**Figure S3 – Fitted individual virus shedding trajectories relative to day of ILI symptom onset (N = 105).** The best-fit model was a Weibull distribution with shape and magnitude parameters modified by age.

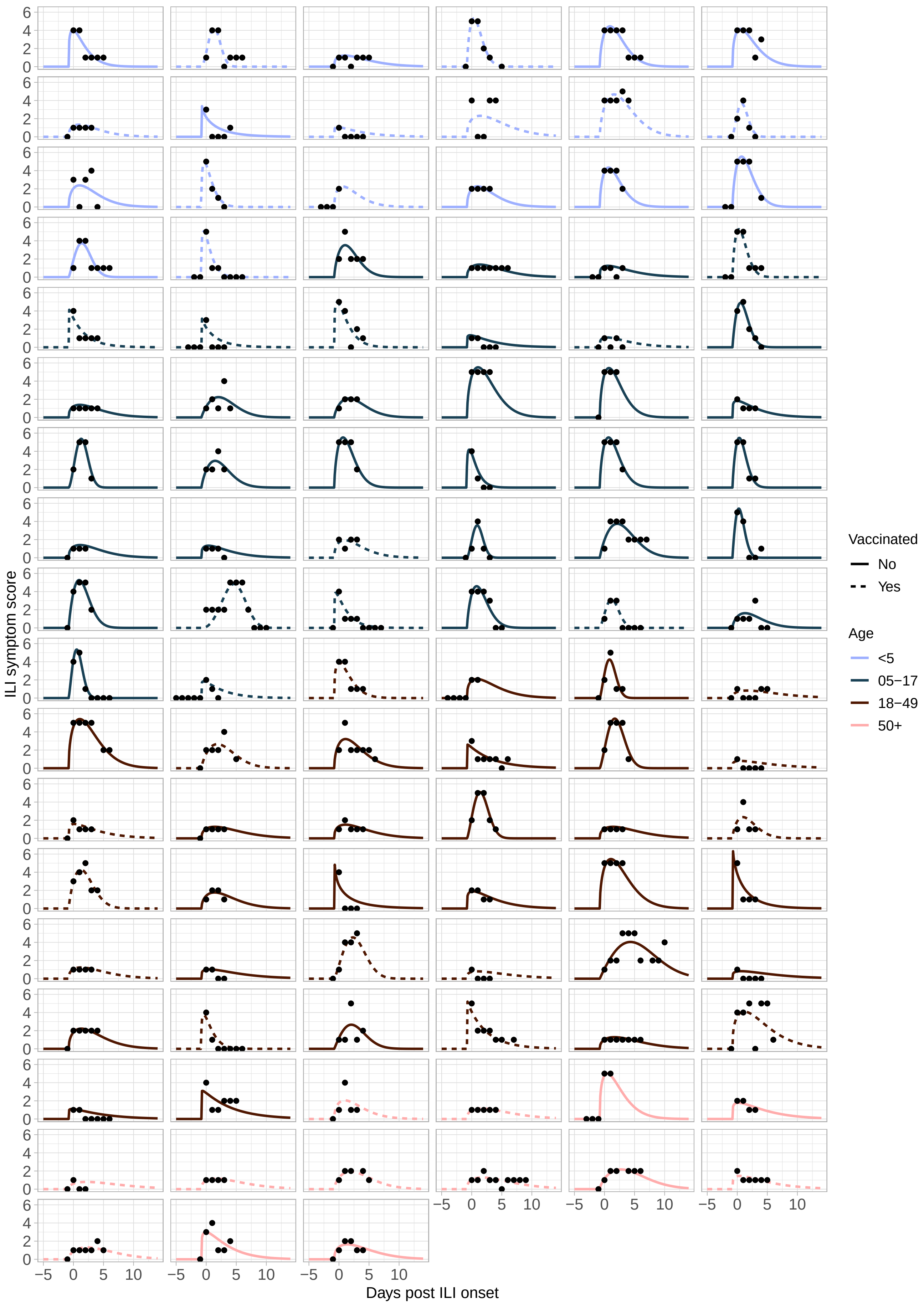


**Figure S4 – Fitted individual $S_{ILI}$ trajectories relative to day of ILI symptom onset (N = 105).** The best-fit model was a Weibull distribution with scale parameter modified by age and magnitude parameter modified by vaccination status.

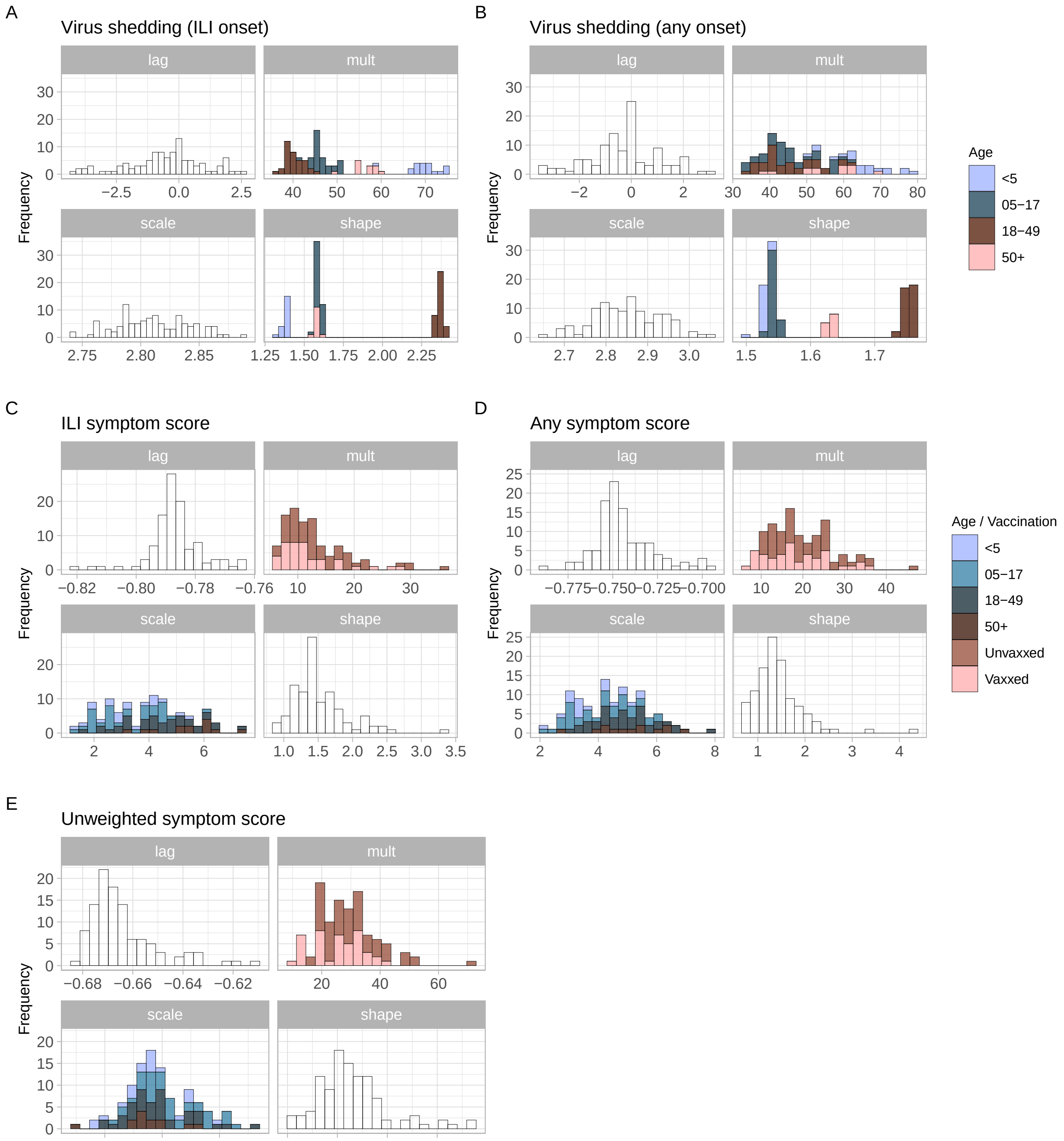


**Figure S5 – Best-fit parameters.** Parameter estimates from the best-fitting models for (A) Ct shedding relative to ILI onset; (B) Ct shedding relative to any symptom onset; (C) S$_{ILI}$ scores; (D) S$_{ANY}$ scores; and (E) S$_{UNW}$ scores. Colors show parameters that were modified by age or vaccination status.

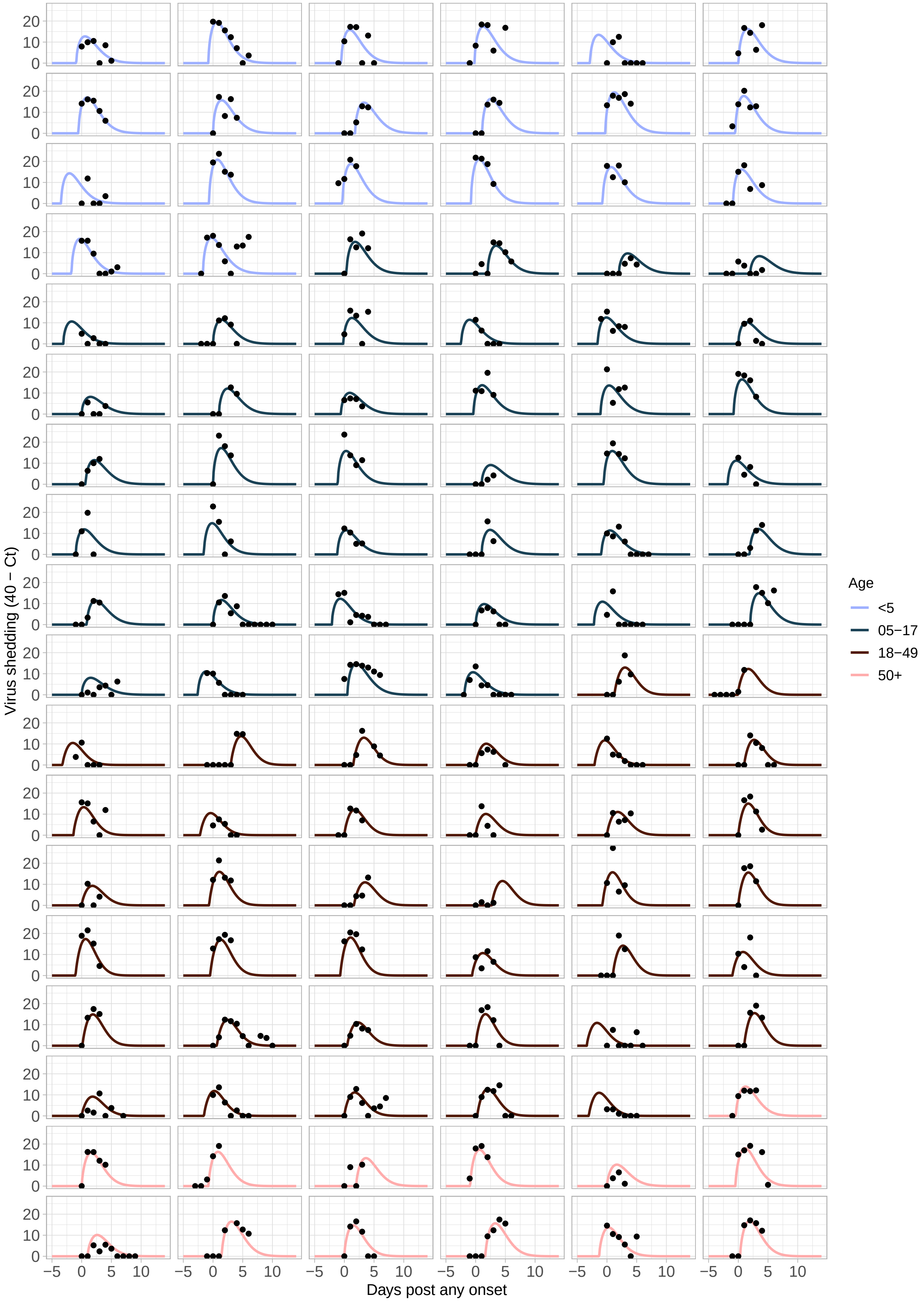


**Figure S6 – Fitted individual virus shedding trajectories relative to day of any symptom onset (N = 108).** The best-fit model was a Weibull distribution with shape and magnitude parameters modified by age.

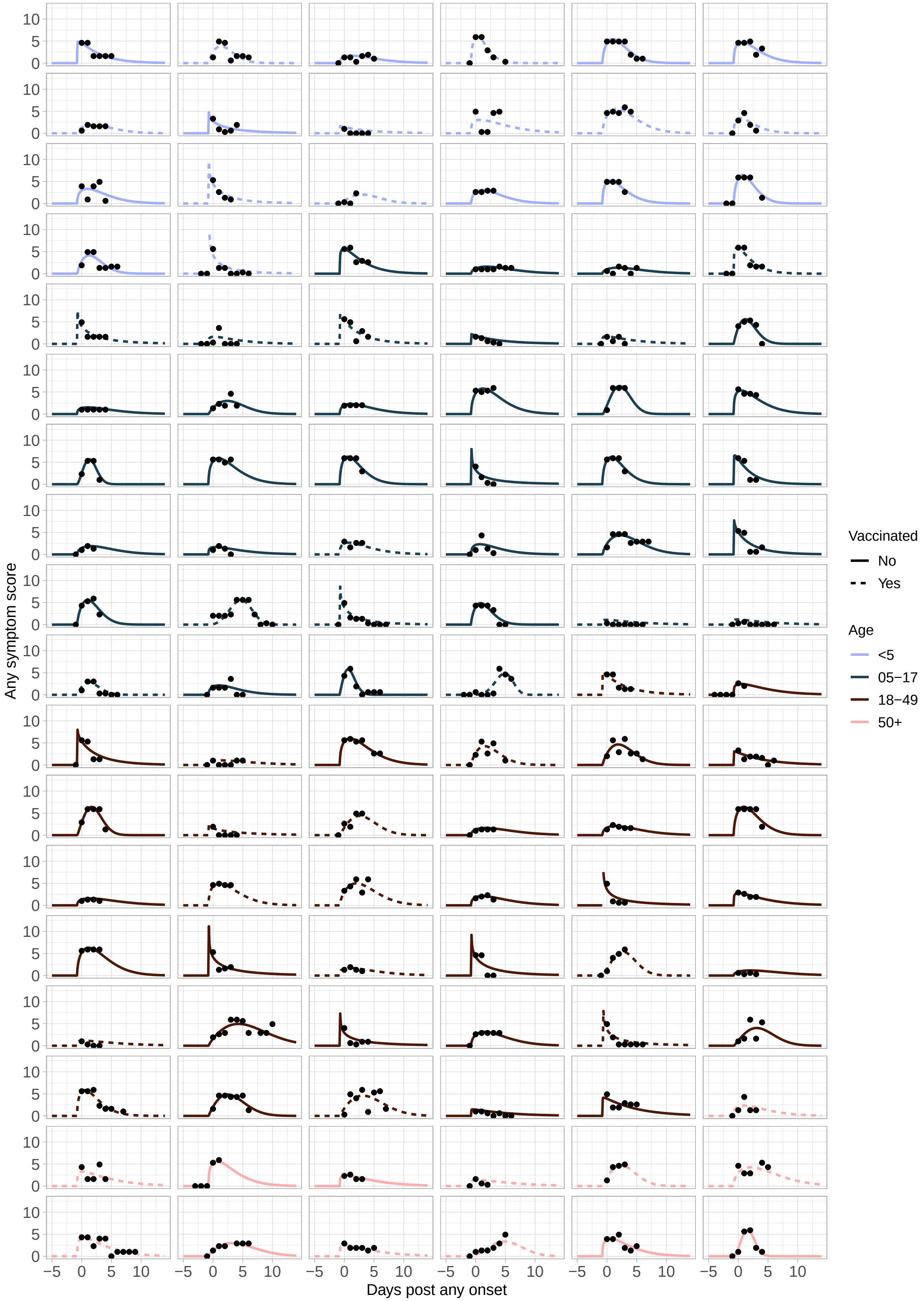


**Figure S7 – Fitted individual $S_{ANY}$ trajectories relative to day of any symptom onset (N = 108).** The best-fit model was a Weibull distribution with scale parameter modified by age and magnitude parameter modified by vaccination status.

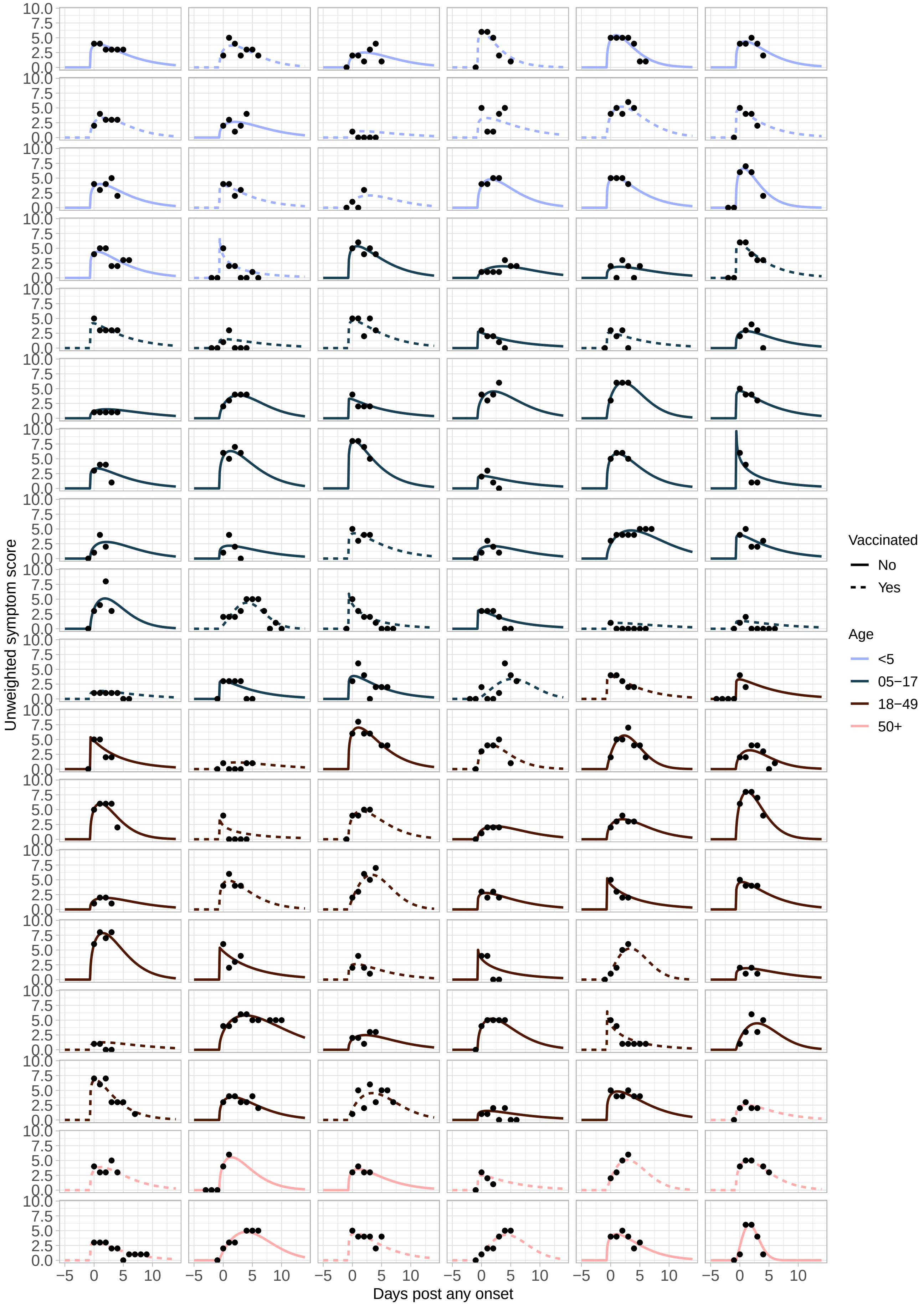


**Figure S8 – Fitted individual $S_{UNW}$ trajectories relative to day of any symptom onset (N = 108).** The best-fit model was a Weibull distribution with scale parameter modified by age and magnitude parameter modified by vaccination status.

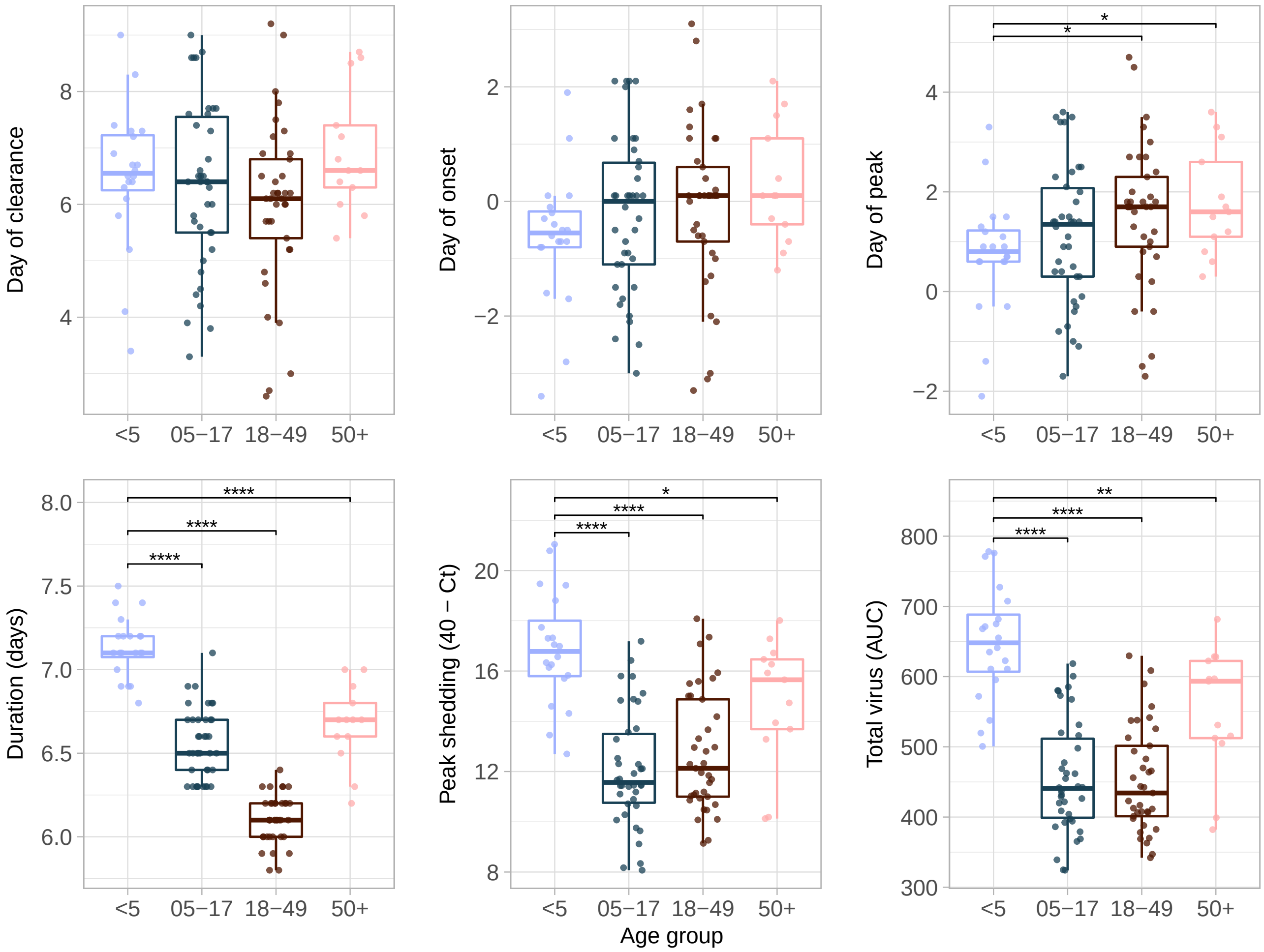


**Figure S9 – Associations between shedding and age using any symptom onset.** Summary metrics top panel from left to right: day of shedding clearance relative to day of any symptom onset; day of shedding onset relative to any symptom onset; day of peak shedding relative to day of any symptom onset. Bottom panel from left to right: duration of shedding in days; peak value of shedding attained (transformed as 40 – Ct); and total virus shed, as measured by the area under the fitted shedding curve. AUC represents the area under the curve. AUC represents the area under the curve. *$p < 0.05$, **$p < 0.01$, ***$p < 0.001$, ****$p < 0.0001$.

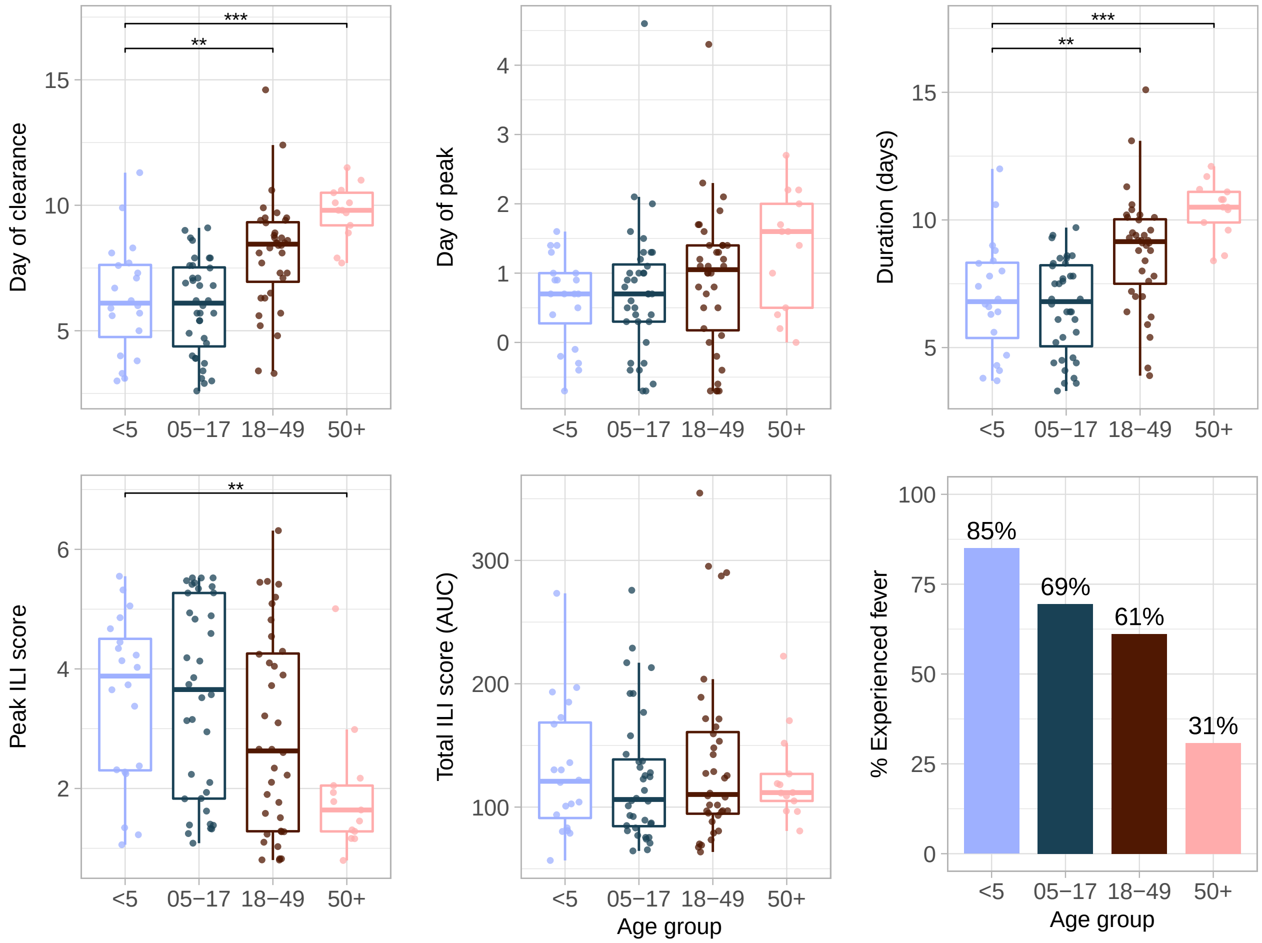


**Figure S10 – Children under 5 have lower peak ILI scores and shorter ILI symptom duration.** Summary metrics top panel from left to right: day of ILI symptom clearance relative to day of ILI onset; day of peak ILI score relative to day of ILI onset; duration of ILI symptoms in days. Bottom panel from left to right: peak ILI score; total ILI score, as measured by the area under the fitted ILI symptom curve; proportion of individuals in each age group experiencing fever (as measured by a fitted ILI score >2). AUC represents the area under the curve. *$p < 0.05$, **$p < 0.01$, ***$p < 0.001$, ****$p < 0.0001$.

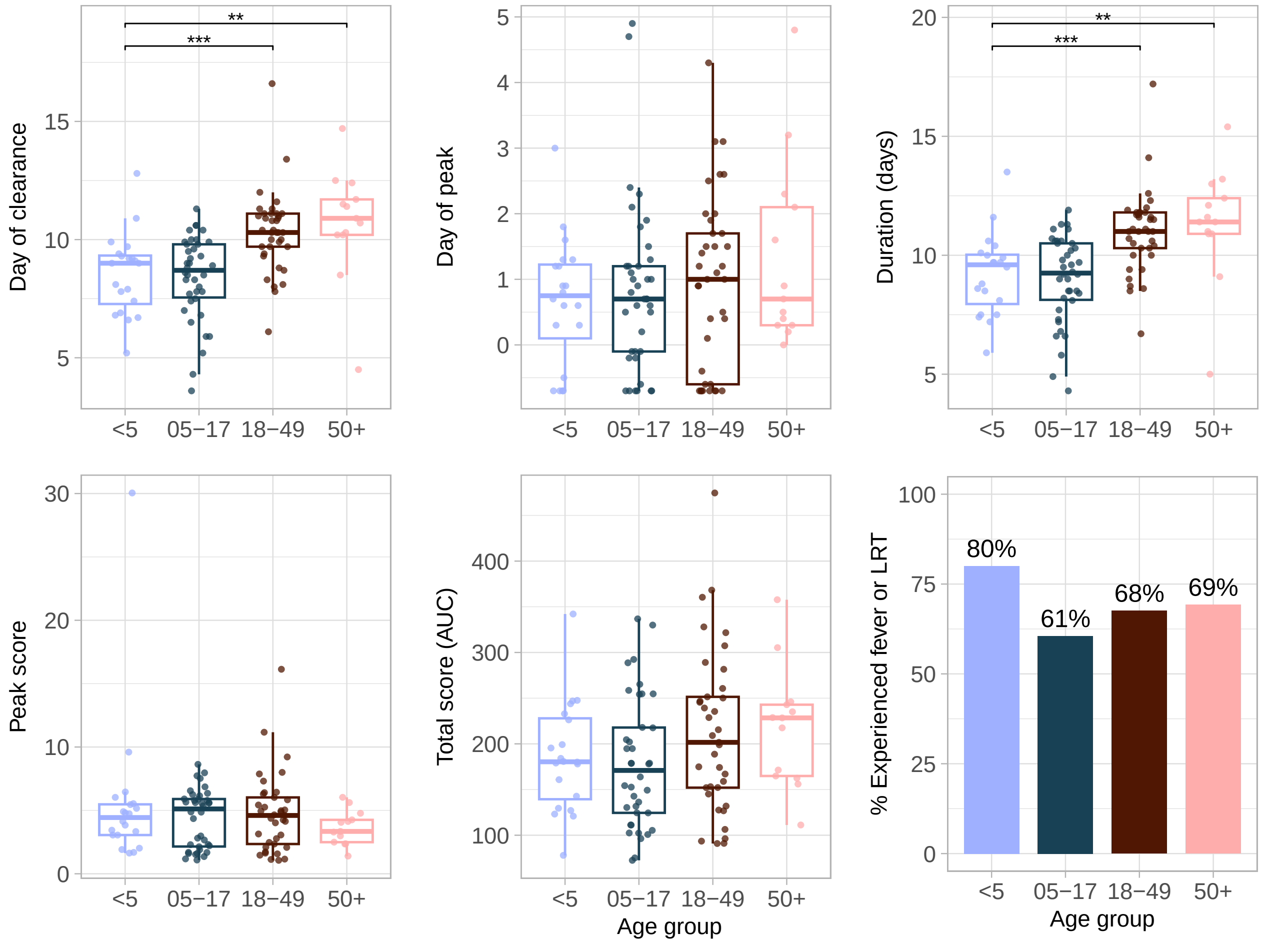


**Figure S11 – $\mathbf{S_{ANY}}$ removes association between peak symptom score and age.** Summary metrics top panel from left to right: day of symptom clearance relative to day of symptom onset; day of peak score relative to day of onset; duration of symptoms in days. Bottom panel from left to right: peak score; total score, as measured by the area under the fitted symptom curve; proportion of individuals in each age group experiencing fever or LRT (as measured by a fitted score $\geq$2.9). AUC represents the area under the curve, and LRT represents symptoms associated with lower respiratory tract infection (i.e., wheezing or shortness of breath). *$p < 0.05$, **$p < 0.01$, ***$p < 0.001$, ****$p < 0.0001$.

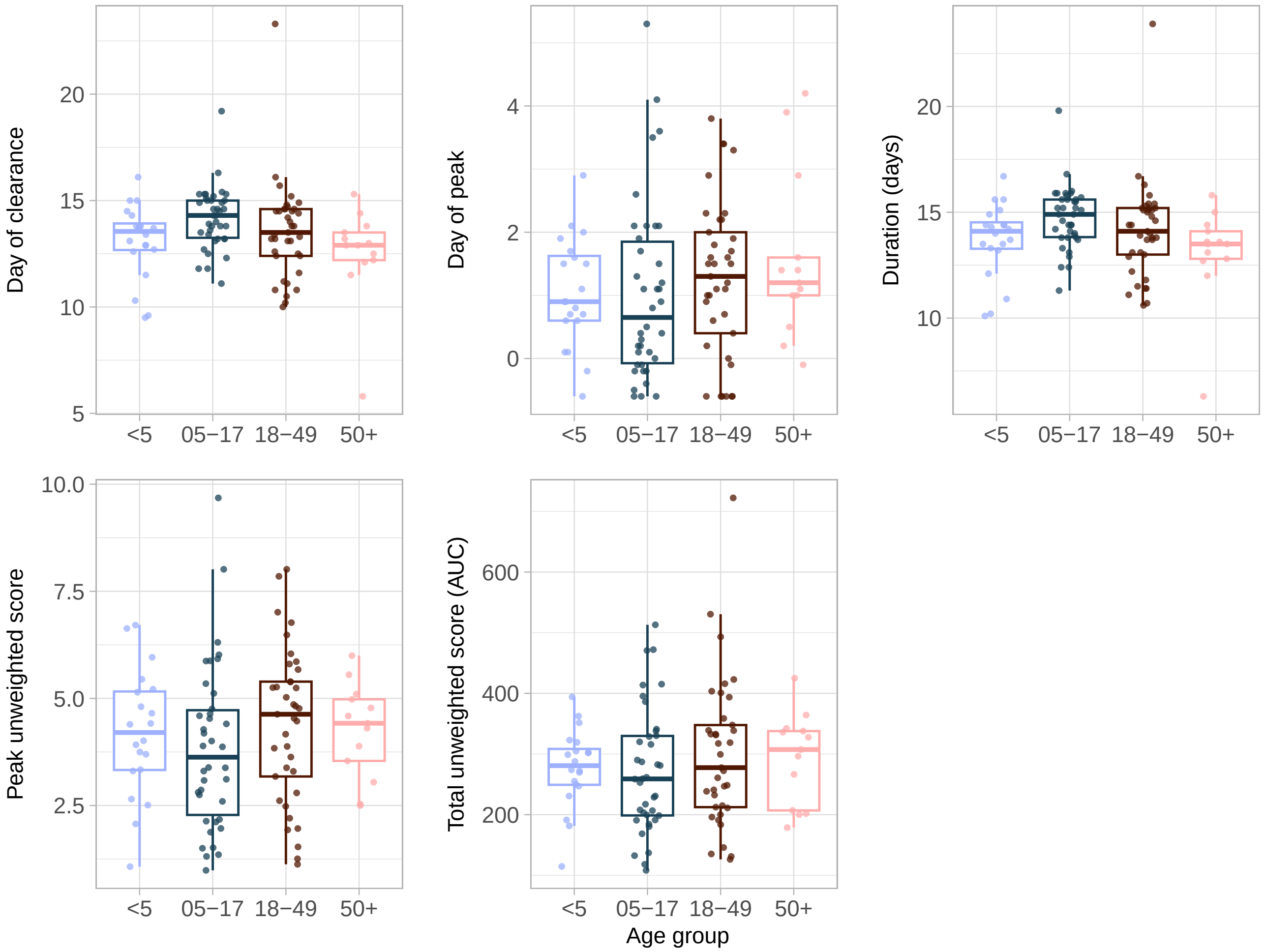


**Figure S12 – $\mathbf{S_{UNW}}$ removes association between peak symptom score and age.** Summary metrics top panel from left to right: day of symptom clearance relative to day of symptom onset; day of peak score relative to day of onset; duration of symptoms in days. Bottom panel from left to right: peak score; total score, as measured by the area under the fitted symptom curve. AUC represents the area under the curve. *$p < 0.05$, **$p < 0.01$, ***$p < 0.001$, ****$p < 0.0001$.

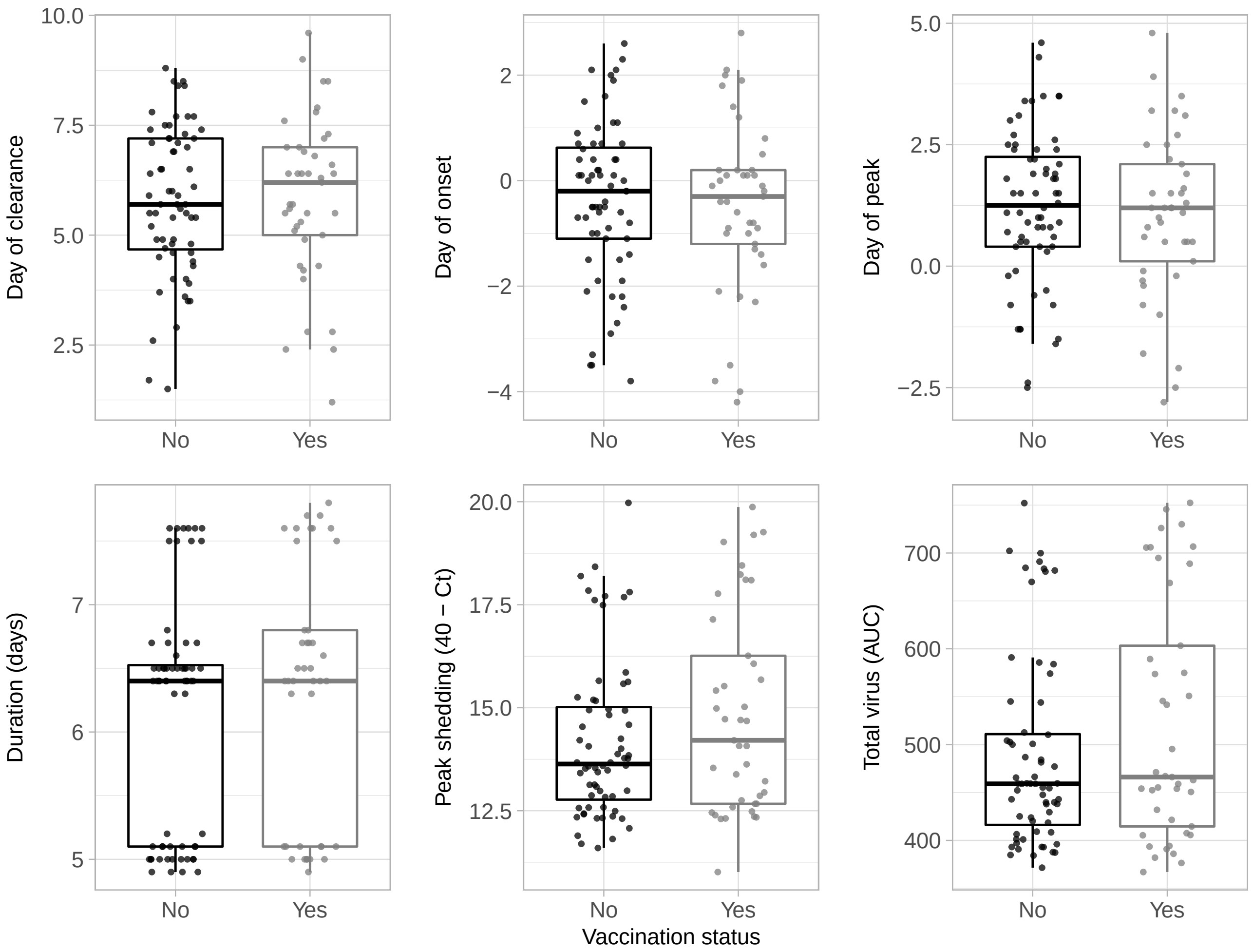


**Figure S13 – No detected associations between shedding and vaccination status.** Summary metrics top panel from left to right: day of shedding clearance relative to day of ILI onset; day of shedding onset relative to ILI symptom onset; day of peak shedding relative to day of ILI onset. Bottom panel from left to right: duration of shedding in days; peak value of shedding attained (transformed as 40 – Ct); and total virus shed, as measured by the area under the fitted shedding curve. AUC represents the area under the curve.

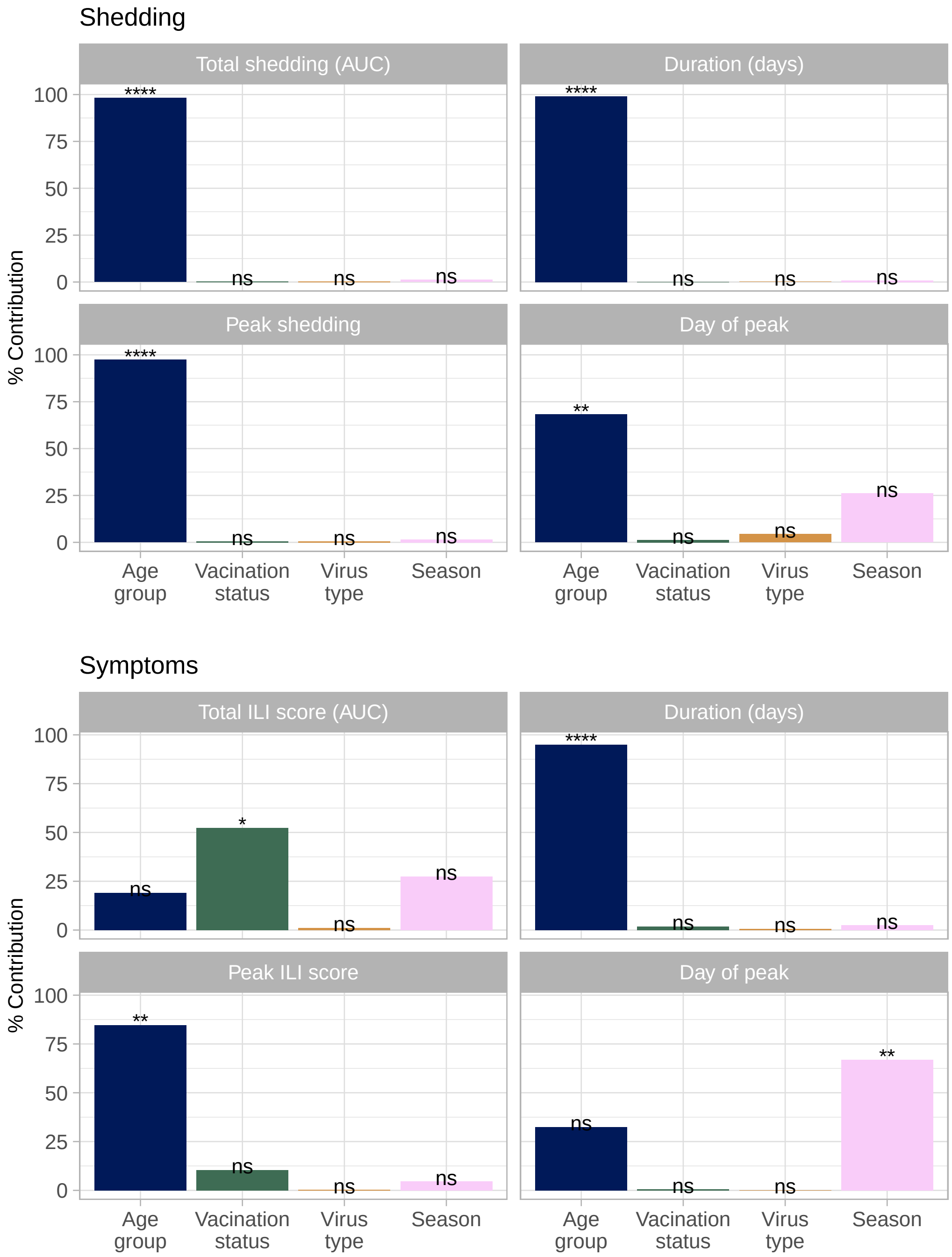


**Figure S14 – Tests of independent covariate contributions using hierarchical partitioning.** Associations between covariates and select summary metrics for shedding (A) or ILI symptom scores (B). The day of peak is relative to days since ILI symptom onset; ILI represents influenza-like-illness; AUC represents area-under-the-curve; and ns represents non-significance. *$p < 0.05$, **$p < 0.01$, ***$p < 0.001$, ****$p < 0.0001$.

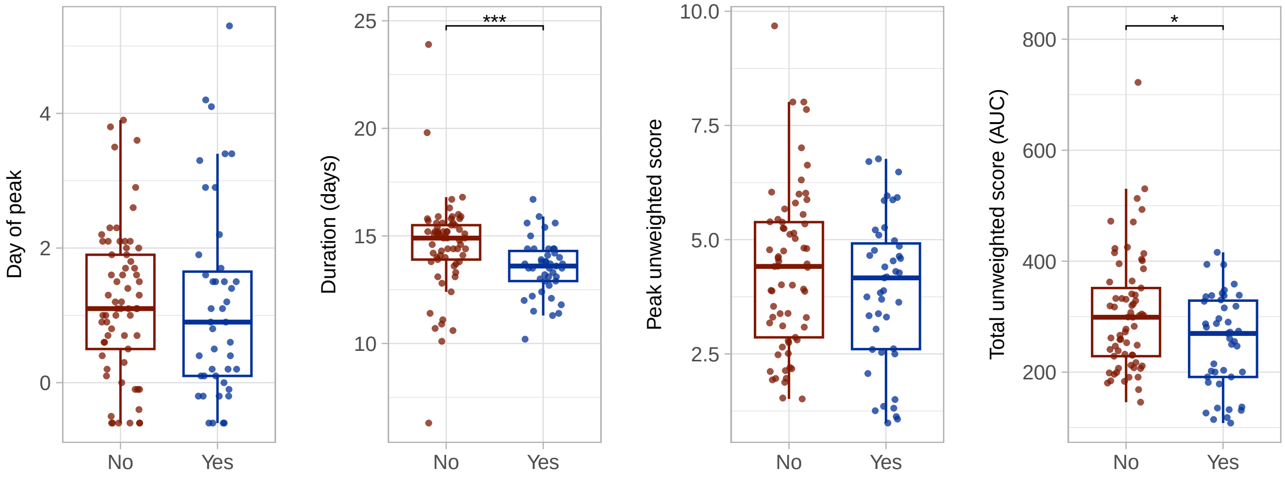


**Figure S15 – Vaccinated individuals experience reduced duration and total unweighted symptom scores.** Days represent days since any symptom onset and AUC represents the area under the curve. *$p < 0.05$, **$p < 0.01$, ***$p < 0.001$, ****$p < 0.0001$.

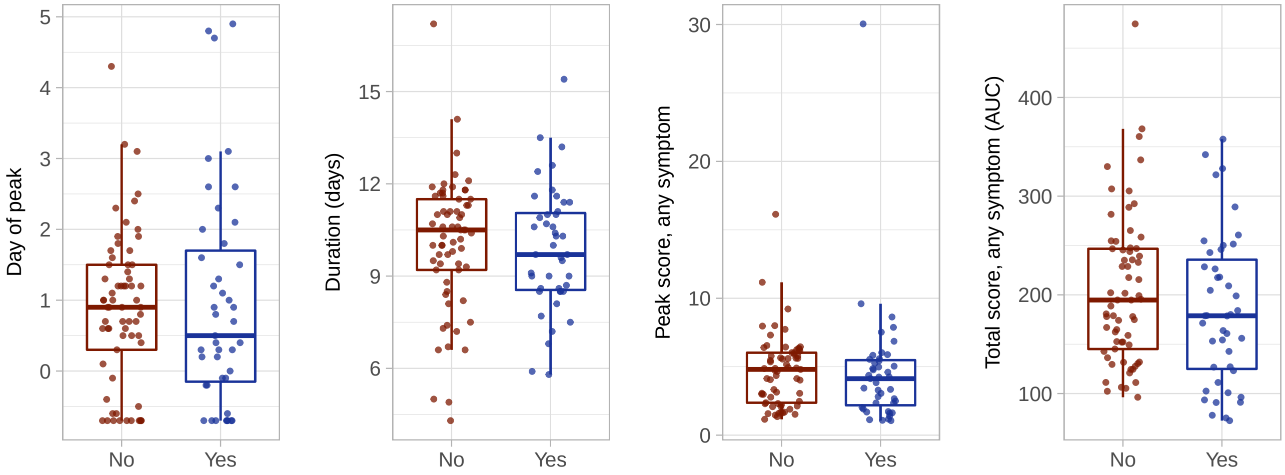


**Figure S16 – No association between vaccination status and $S_{\text{ANY}}$.** Days represent days since any symptom onset and AUC represents the area under the curve.

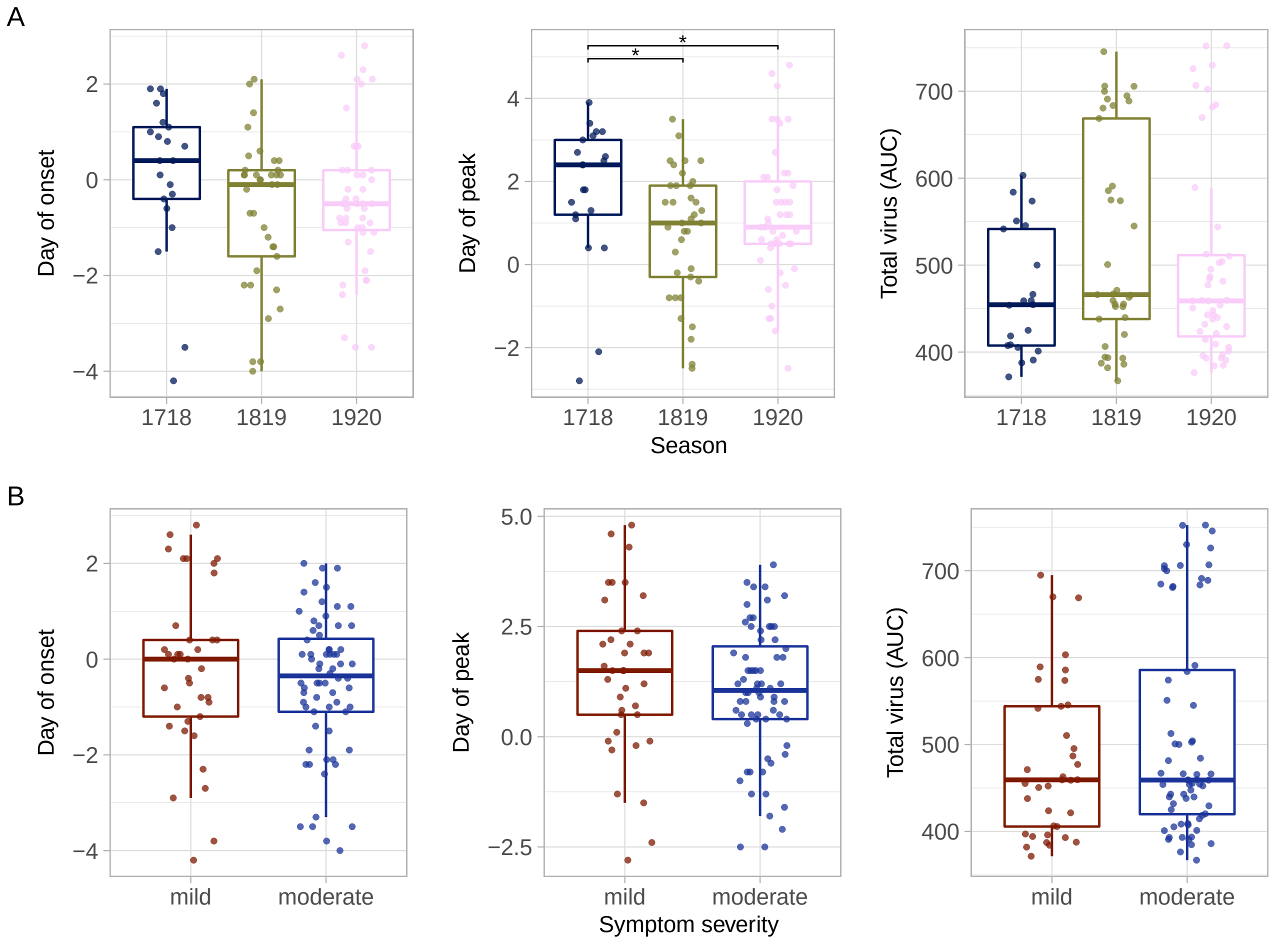


**Figure S17 – Additional associations between virus shedding and season (A) or symptom severity category (B).** Shown are associations with day of shedding onset (left); day of peak shedding (middle); and total virus shed (right). Individuals with peak ILI score $> 2$ are classified as experiencing 'moderate' symptoms; all others are classified as experiencing 'mild' symptoms. Days represent days since ILI symptom onset and AUC represents the area under the curve. *$p < 0.05$, **$p < 0.01$, ***$p < 0.001$, ****$p < 0.0001$.

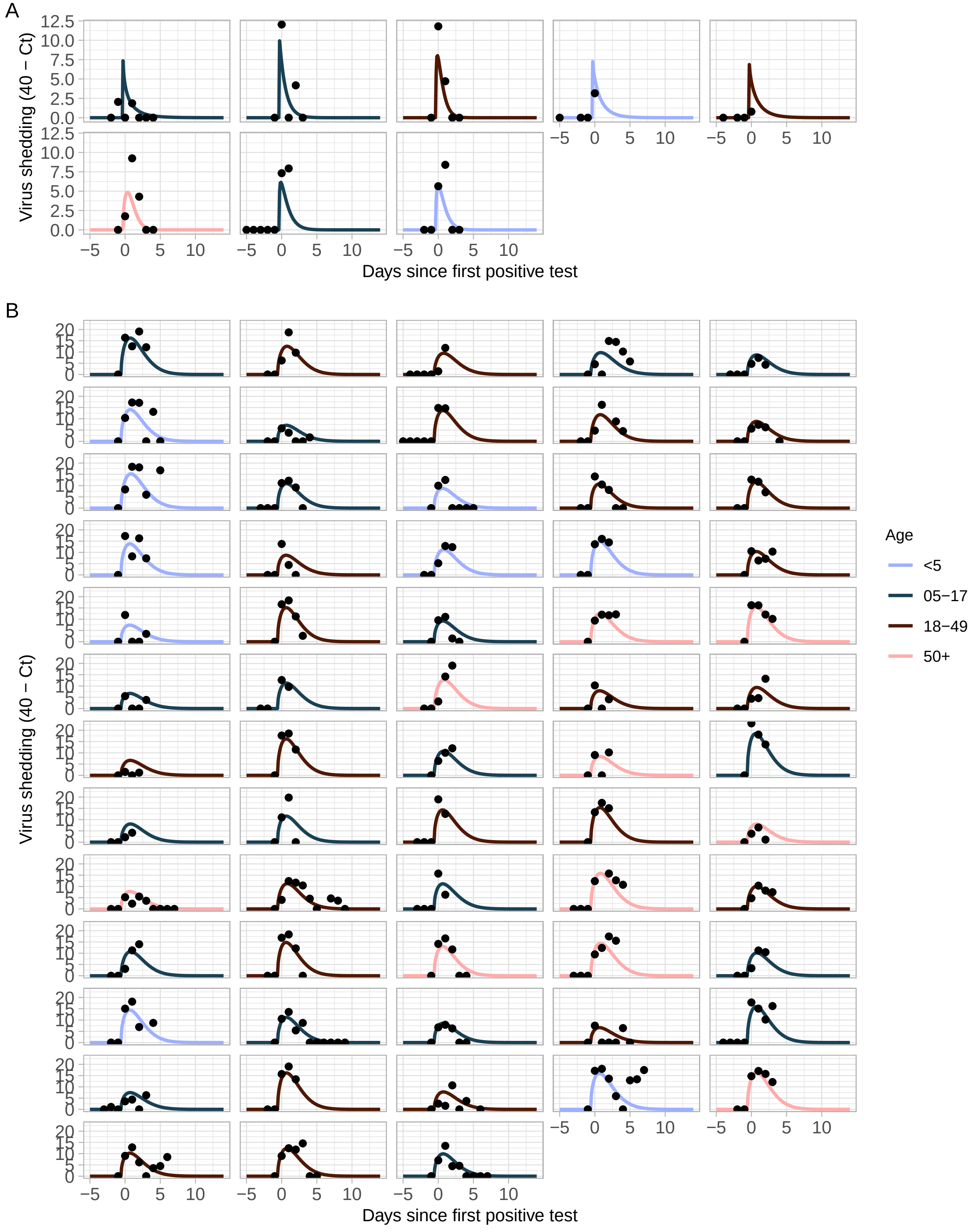


**Figure S18 – Fitted individual virus shedding trajectories relative to day of first positive test.** Fits for asymptomatic (A) and symptomatic (B) individuals with incident infection from the best-fitting Weibull distribution. Model parameters were not modified by any covariates; age group is shown for reference only.

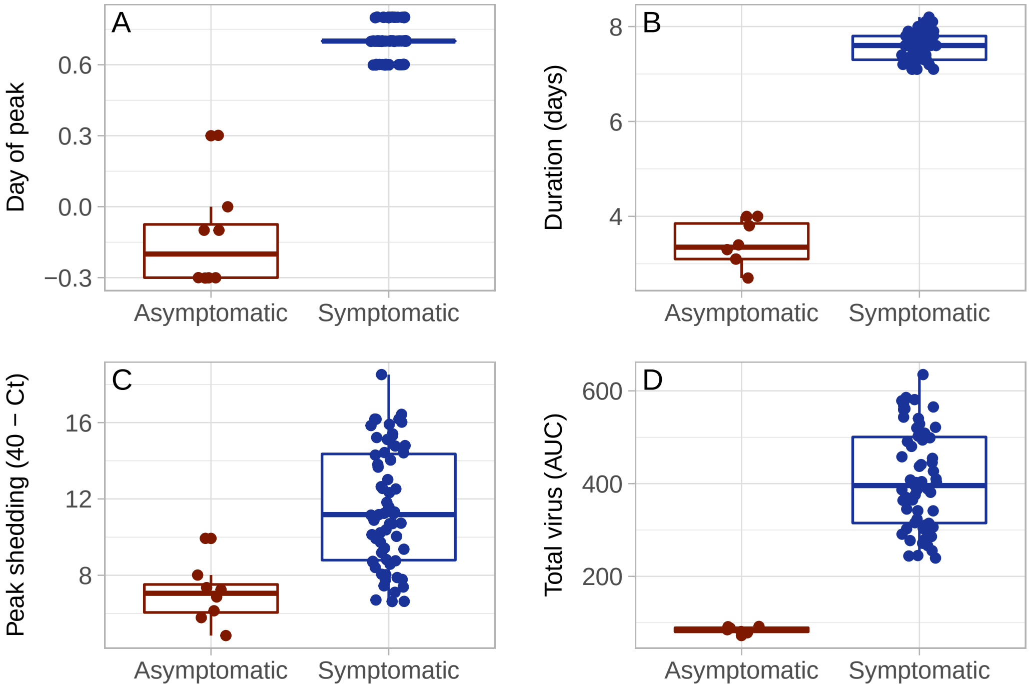


**Figure S19 – Summary metrics for asymptomatic and symptomatic individuals with incident infection.** Shown are: (A) day of peak shedding clearance relative to day of first positive test; (B) duration of shedding in days; (C) Peak value of shedding attained (transformed as 40 – Ct); and (D) total virus shed, as measured by the area under the fitted shedding curve (AUC).

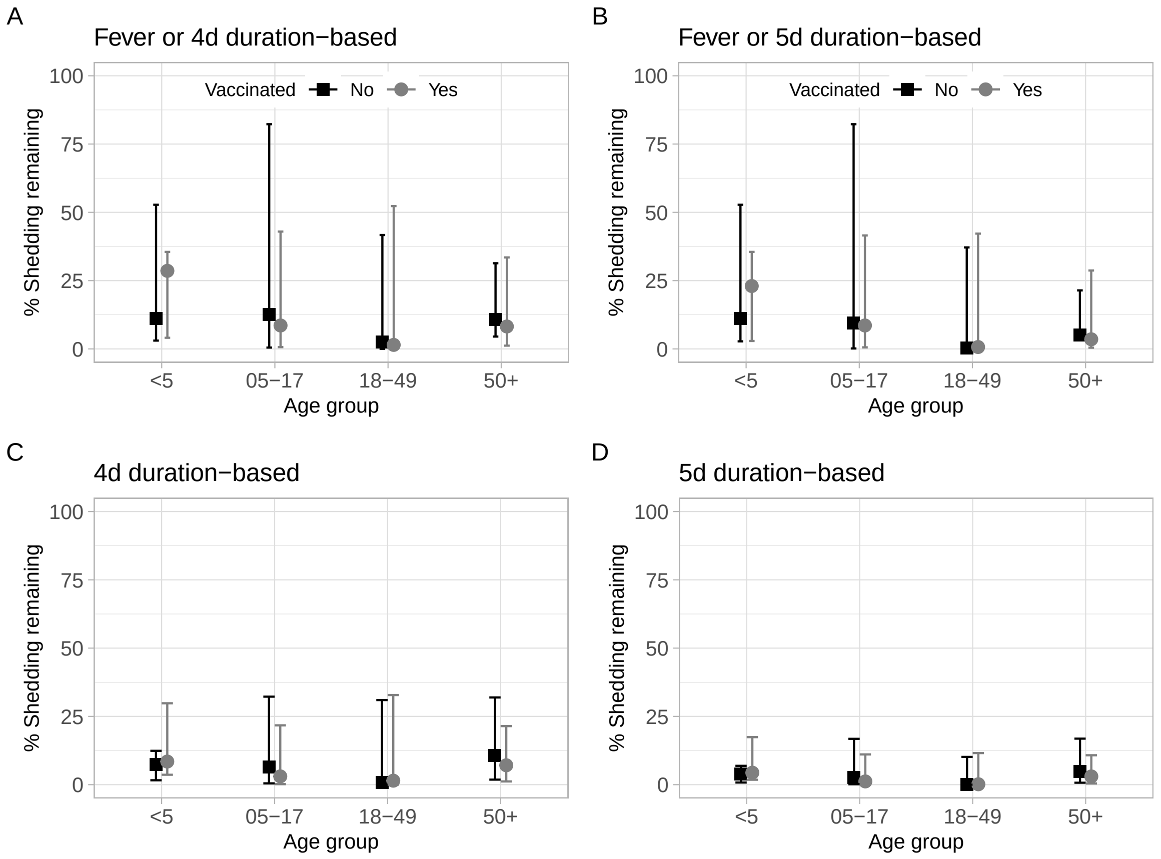


**Figure S20 – Duration-based isolation guidelines are effective in reducing shedding from symptomatic individuals.** (A, B) Estimates of shedding assuming current isolation guidance is followed. Individuals who do not experience fever isolate for either 4 days (A) or 5 days post ILI symptom onset. (C,D) Estimates of the percentage shedding assuming all individuals follow duration-based isolation guidance, regardless of symptoms, for 4 (C) or 5 (D) days. Estimates are stratified by age and vaccination status. Points represent the median within each stratification and error bars are the 90th percentiles.
